# Genome-wide associations spanning 194 in-hospital drug dosage change phenotypes highlight diverse genetic backgrounds in concurrent drug therapy

**DOI:** 10.1101/2025.02.04.25321575

**Authors:** Alexander Pil Henriksen, Cristina Leal Rodríguez, Hannah Currant, Ioannis Louloudis, Jorge Hernansanz Biel, Maria Herrero-Zazo, Ewan Birney, Thomas Folkmann Hansen, Gianluca Mazzoni, Amalie Dahl Haue, Henning Bundgaard, Christian Erikstrup, Khoa Manh Dinh, Liam Quinn, Mie Topholm Bruun, Henrik Hjalgrim, Erik Sørensen, Christina Mikkelsen, Michael Schwinn, Ole Birger Vestager Pedersen, Henrik Ullum, Sisse Rye Ostrowski, DBDS Genomic Consortium, Karina Banasik, Søren Brunak

## Abstract

As populations get older and medicine consumption rises, the rate of concurrent drug use and polypharmacy among patients is increasing. Polypharmacy is known to complicate therapy and increase the risk of drug-drug interactions, the individuality of which remain largely unexplored. Here, we perform a series of genome-wide association studies to identify variants associated with dosage changes during episodes of concurrent drug therapy. We extracted in-hospital drug prescription records from 847,537 patients in a population-wide Danish hospital cohort. Using imputed genotype data from the Copenhagen Hospital Biobank and the Danish Blood Donor Study we then performed a series of genome-wide association analyses across 194 drug pair phenotypes fulfilling selection criteria. We identified 51 genome-wide significant (p < 5e-08) loci, 49 so far unreported in any genome-wide association studies, associated with dosage changes across 42 different drug pair phenotypes. 49 of the identified loci were unique to the respective drug pairs. Through annotation of the identified loci, expression quantitative trait loci analyses, and gene-based tests we found links to 57 distinct genes, several of which have previously been associated with disease.

This study identifies genes that may modulate response to drug therapy in the context of polypharmacy. Our findings reveal distinct patterns of genetic variation across different drug pairs, suggesting a diverse set of genes involved in drug efficacy and drug response. This study may give a better understanding of the individuality of such mechanisms and may aid the development personalized treatment approaches.

## Background

Globally medication consumption is on the rise^1,2^. There is a notable increase in patients receiving several medications concurrently, particularly among the elderly and others with multimorbidities ^3–6^. It is well established that polypharmacy, commonly defined as taking five or more medications, is associated with adverse drug events (ADEs) as well as higher healthcare costs^7–9^. Use of multiple drugs may also lead to more subtle therapeutic changes such as changes in drug dosage. In a Danish cohort of more than a million patients Leal et al. recently identified 3,993 unique drug pairs, many with known drug-drug interactions (DDIs), which were significantly associated with an increased probability of dosage adjustments^10^. DDIs are well known to be enriched in certain types of drug therapy and have also been shown to contribute to increased rates of hospital admissions, prolonged hospital stays, and more specific clinical outcomes such as bleeding and toxicity^9,11–13^.

The importance of pharmacogenetics, the effect of individual genetic variation on drug response, is becoming increasingly clear^14^. For example, deleterious variants in drug-metabolizing genes may lead to drug concentrations far outside the therapeutic range. Such drug-gene interactions (DGIs) have been shown to be relevant for up to 24% of patients receiving drug therapy and the added effects of drug-drug-gene interactions from concurrent medications may raise this level even further^15,16^.

Despite the wealth of documented drug-drug and drug-gene interactions, the underlying biological mechanisms for many interactions remain elusive. Existing databases such as DrugBank, NSIDES, and PharmGKB offer valuable insights but lack full coverage, leaving a substantial portion of DGIs unexplored, especially in the context of polypharmacy^17–19^. While databases may list hundreds or thousands of DGIs and DDIs, many interactions are without known biological mechanisms and without clinical evidence.

This study aims to address this gap by investigating the genetic underpinnings of dosage adjustments resulting from concurrent drug use. While some dosage changes can reflect standard clinical practice, others may reflect changes in drug absorption, distribution, metabolism, or excretion, processes which may be influenced by interacting drugs and/or genetic factors. In a population of 133,467 genotyped Danish patients covering 436,367 admissions between 2009 and 2016, we perform genome-wide association study (GWAS) analyses to investigate if genetic variants are associated with drug dosage changes in concurrent drug use. We present GWAS results of concurrent drug use phenotypes for 194 drug pairs. The results highlight several genes that may help explain both intended and unintended effects of concurrent drug use.

## Methods

### Data sources

In brief, in-patient drug administration data were gathered from electronic healthcare records of more than one million patients admitted to hospitals in Eastern Denmark between 2008 and 2016. We used medication data covering 185 million treatment episodes described in detail in Leal et al.^10^. These data were organized into drug pair treatment episodes in which individual patients are prescribed two or more drugs at the same time. Each treatment episode included an index drug and a codrug and the daily dose of the index drug was tracked during periods of concurrent prescription as well as during monotherapy. The analysis identified 3,993 drug pairs for which taking the pair of drugs concurrently significantly increased the probability of dosage changes of the index drug when compared to taking the index drug as monotherapy. These pairs are, as in Leal et al.^10^, referred to as “dosage adjusted drug pairs”.

Genotype data was retrieved from the Copenhagen Hospital Biobank (CHB)^20^, a research biobank which contains samples obtained during workup on hospitalized patients and outpatients at hospitals in the Capital Region of Denmark^20^ and from the Danish Blood Donor Study, a large prospective cohort of blood donors initiated in 2010^21^. Our analyses were limited to the CHB + DBDS Oral cohort for which we had been granted access by Danish authorities. The Oral cohort contains 375,217 patients of which 177,622 had hospital drug records containing any of the 3,993 dosage adjusted drug pairs. Genotyping was performed at deCODE genetics using the Illumina Infinium Global Screening Array. Standard quality control measures were applied and imputation was performed using a North-Western European reference as described elsewhere^22^. All genotype data was aligned with the GRCh38 Human Reference Genome build.

For a follow-up analysis, primary care prescription records were retrieved from the Danish National Prescription Registry (DNPR), which contains all prescription drugs collected at Danish community pharmacies^23^. DNPR contains individual-level records for prescriptions redeemed since 1994 and includes information such as date of redemption, drug, brand, dose, and pack size.

### Drug pair selection

177,622 patients had hospital drug records containing any of the 3,993 dosage adjusted drug pairs as well as accessible genotype information (Table S1). We limited our analysis to the most densely populated, well-mixed population within the overall dataset using visual inspection in the principal component PC1-PC2 space; a population which largely aligns with those who identify as Danish / Northern European. This left 133,467 patients, covering 436,367 separate admissions and 3,950,941 treatment episodes. To reduce the number of drug pairs, our analysis was limited to those episodes in which a patient had received more than a single prescription of the index drug. We further excluded drug pairs that included Ibuprofen or Paracetamol as these medications are readily prescribed in hospitals and are often given as “pro re nata” (i.e. by the patient’s needs), making it difficult to know to what extent the patient has taken the drug. When patients had been prescribed a drug pair during multiple separate hospital admissions, we limited our analysis to the first admission to avoid confounding from later clinical intervention. These exclusions left us with 3,714 drug pairs. Finally, we used the CaTS Power Calculator to filter drug pairs to only include those which we had a sufficient sample size to detect a genotype relative risk of 1.2 with 80% power^24^. This left us with 194 drug pairs covering 332,904 admissions and 117,814 patients (Figure S1, Table S2). The included drug pairs were prescribed to between 1,345 and 26,193 patients.

### Genetic analyses

The drug pair selection left us with 194 drug pair phenotypes for which we tested genetic association with dosage changes of the index drug. We encoded dosage change as a binary phenotype with “Change” being when a patient had received a pair of drugs during a hospital admission and had changed the dosage of the index drug between two prescriptions. “No change” controls had also received the pair of drugs but had not changed dosage of the index drug between two prescriptions. The sample size of the case population (i.e. patients experiencing a “Change”) ranged from 998 to 18,873 across drug pair phenotypes (Table S2). We performed GWAS for each of the drug pair phenotypes using an additive logistic model with REGENIE v. 3.1^25^. Model covariates included, sex, year of birth, age at first administration of drug pair, age squared, hospital of admission, genotyping chip, and the first six genetic PCs. The number of PCs was chosen by including the top PCs until the explained variance plateaued (Figure S2). After each GWAS, we removed variants with INFO scores lower than 0.9, multiallelic variants, and variants which did not pass basic quality control in the overall cohort (Minor Allele Frequency > 0.01, genotype missingness < 0.1, Hardy-Weinberg equilibrium exact test p-value < 10^−15^).

GCTA Conditional and Joint Analysis (COJO) as provided through GCTA^26,27^ was used to select significantly associated independent loci which were more than 10Mb apart.

Significant variants were annotated using ANNOVAR^28^ and OpenTargets^29^. Where inconsistencies were observed we kept the OpenTargets annotation. Linkage disequilibrium scoring was done using LDSC^30^ and gene-based testing was done using fastBAT^31^ through GCTA^27^. We tested variant effects on dosage change directionality using linear regression in R v.4.3.3 with all the same covariates as used in the original GWASs. We used the eQTL Catalogue API^32^ to perform eQTL mapping. Additionally, FUMA was used to perform gene set enrichment analysis. FUMA’s GENE2FUNC pipeline was run using default settings, submitting only genes identified by location, gene-based testing or eQTLs for each drug pair phenotype. PheWAS results for individual variants were extracted from the OpenTargets database^29^. We corrected for multiple testing using the Bonferroni method counting the number of datasets included (p < 1.25e-05).

### Correlation between drug pair phenotype genetic effects

Among the 194 drug pair phenotypes tested for genetic associations many shared index- or codrugs. We speculated whether the genetic background driving dosage change in one drug pair could be similar to that of another drug pair with the same index drug. To calculate the genetic correlation between drug pair phenotypes we created a variant set consisting of all variants that had a p-value of 5e-7 or lower in any of the 194 performed GWASs. The effect sizes of the variants in this variant set were then compared and a Pearson correlation was calculated between each combination of two drug pair phenotypes.

### Variant effects on primary care prescription patterns

We extracted prescription records from the DNPR (1994–2022) for the 375,217 individuals from the Oral cohort. 374,406 (99%) individuals had available records in DNPR. For each variant of interest, patients were stratified by genotype (0, 1, 2) and prescription records for the two drugs relating to that variant was extracted. Patients included in the GWAS analyses originally identifying each of the 51 genome-wide significant variants were excluded from the analysis. We separated prescription records into treatment episodes where patients had either 1) received the index drug from a drug pair or 2) received both the index drug and codrug in a drug pair within 30 days of each other. We performed linear regression to assess associations between variant genotype and two phenotypes relating to the index drug: 1) number of redeemed prescriptions, and 2) mean dose of redeemed drug. We included sex, date of birth, and age at first prescription as covariates. For both phenotypes we excluded the 1% most extreme values (1% top values). We performed all statistical analyses using R v4.3.3.

## Results

### Discovery cohort demographics

We started with 133,467 patients who had been prescribed one of 3,993 dosage adjusted drug pairs during hospital admission. The index drugs prescribed to most patients were those belonging to anatomical therapeutic chemical (ATC) classification groups N02 (Analgesics, 53.1%), J01 (Antibacterials for systemic use, 51.7%), and B01 (Antithrombotic agents, 45.2%). The most common index drugs were potassium chloride (a common salt, 40.7 % of admissions), morphine (an analgesic, 38.2 % of admissions), and cefuroxime (an antibiotic, 34.5 % of admissions). We selected 194 drug pairs with sufficient sample size to run a GWAS (Figure 1A). These drug pairs had been prescribed to between 1,345 and 26,193 patients in total with the fraction of patients experiencing a dosage change (cases) varying between 26.8% and 87.7%. Of the selected 194 pairs a 60.8% (118) contained drugs from different ATC groups while the pairs with drugs from the same ATC groups were mainly from groups B01 and J01 (Figure 1B). The median number of patients for the 194 drug pairs was 4,529 (sd = 4,114), with 59% of pairs (114) being prescribed to more than 4,000 patients (Figure 1C). Drug pairs with index drugs in ATC group J (Antiinfectives for systemic use) had generally been prescribed to fewest patients (median = 3,388 patients) while those with index drugs in ATC group N (Nervous system) had generally been prescribed to most patients (median = 7,860 patients).

**Figure 1:**
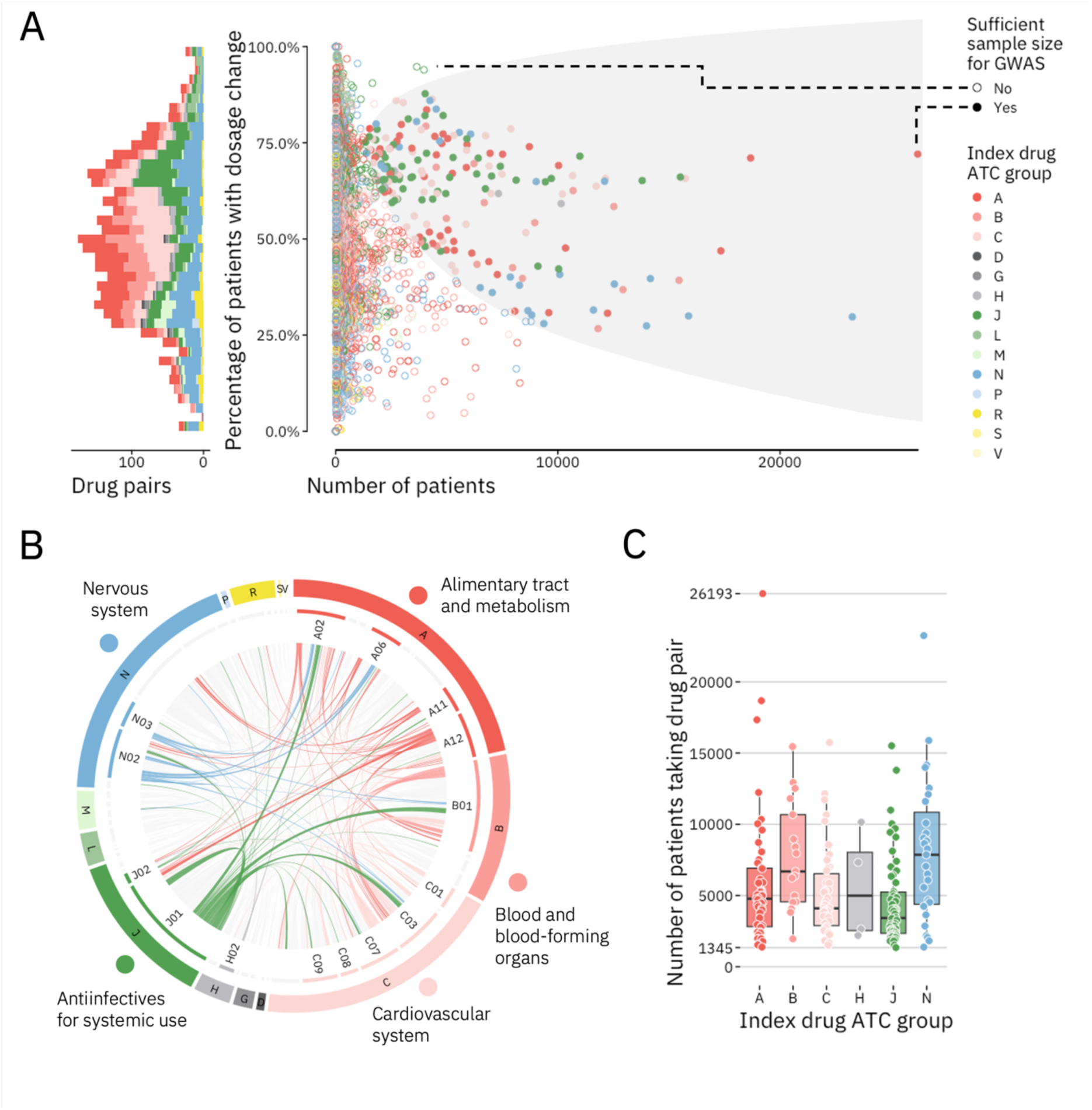
Overview of the selected drug pair phenotypes. (A) Patient counts, and dosage change rates for 3,714 relevant drug pairs. Left: A stacked histogram of the drug pairs color-coded by the ATC code of their index drug. Right: Scatter plot showing the number of patients and dosage change rate of each included drug pair. Solid circles are drug pairs administered to enough patients to be selected for GWAS. Left: A stacked histogram of the drug pairs color-coded by the ATC code of their index drug. (B) Circos plot of 3,714 relevant drug pairs stratified by ATC drug groups. Links connect the index drug and codrug for each drug pair. Drug pairs with sufficient sample size for GWAS are colored. Links are colored as in A. (C) Number of patients with the selected drug pair phenotypes. Each dot shows the number of patients receiving that drug pair. Boxplots are split by index drug and colored as in A.

### GWASs identify 51 variants associated with dosage changes

We identified 51 genome-wide significant (p < 5e-8) loci associated with dosage changes in 42 of the drug pairs (Figure 2, Table 1, for full variant information see Table S3. Manhattan- and QQ plots are available in Figure S5). No variants reached the significance level required by multiple test correction with Bonferroni (p < 5e-8 / 194). There was no overlap in the lead variants between drug pair phenotypes. However, rs2511771 (tinzaparin + potassium chloride) and rs56255127 (potassium chloride + magnesium) were located within 1 mbp of each other and both mapped to the *NTM* gene (OpenTargets annotation).

**Figure 2:**
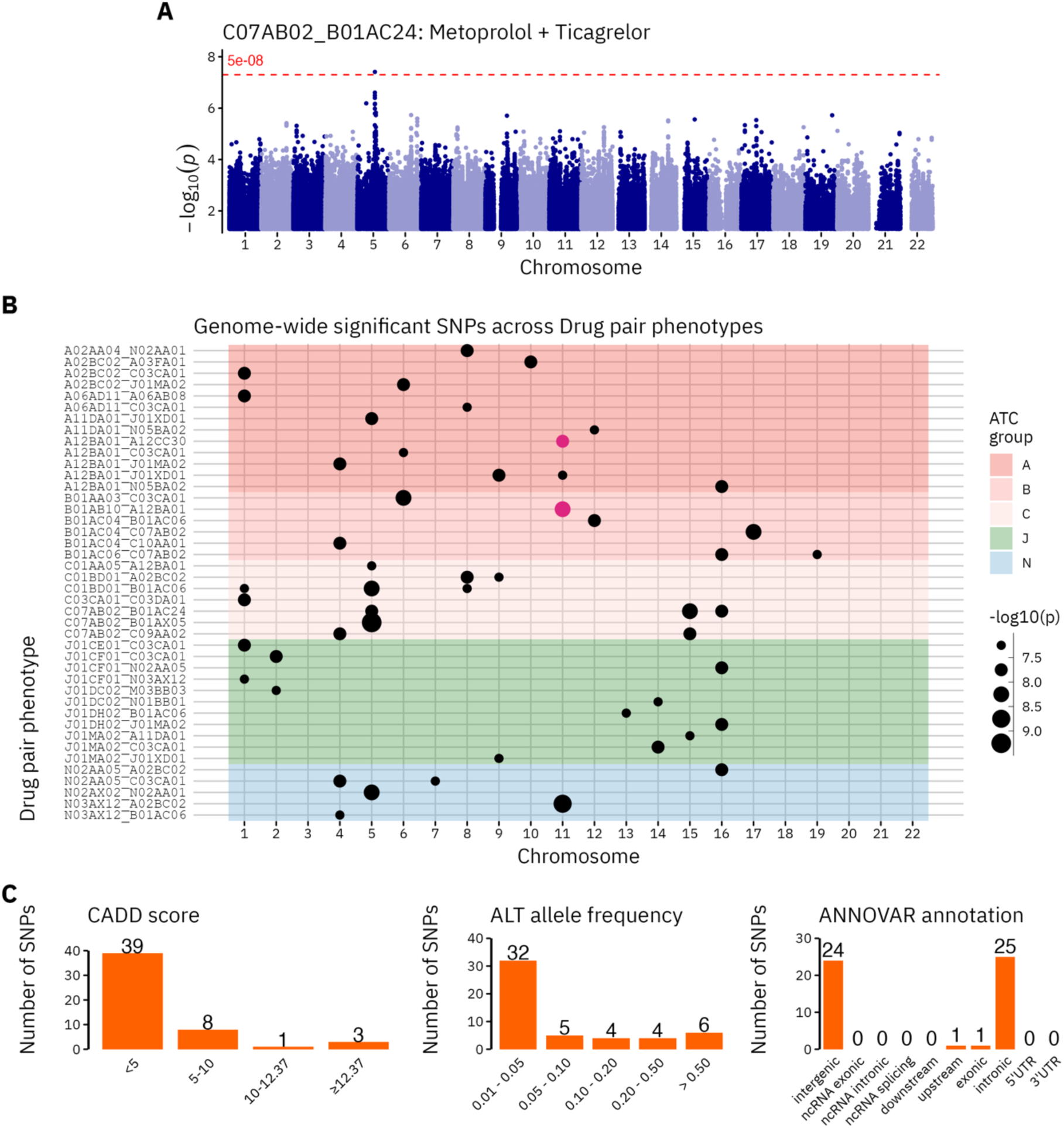
Significant SNP results from the 194 drug pair phenotype GWASs (42 with significant hits). (A). Manhattan plot of the Metoprolol + Ticagrelor drug pair with one significant locus. (B). Discrete scatterplot of all significantly associated SNPs for each drug pair phenotype. The dot size indicates the significance level. Purple dots in the same column indicate pairs of SNPs that are within 1 mbp of each other. The background color indicates the ATC group of the index drug in the drug pair. (C). Distribution of SNPs across CADD scores, allele frequencies and ANNOVAR functional annotations. CADD scores predict the pathogenicity of a SNP with values above 12.37 being considered potentially pathogenic. ANNOVAR annotation categories identify the position of variants relative to nearby genes and their associated function.

**Table 1:**
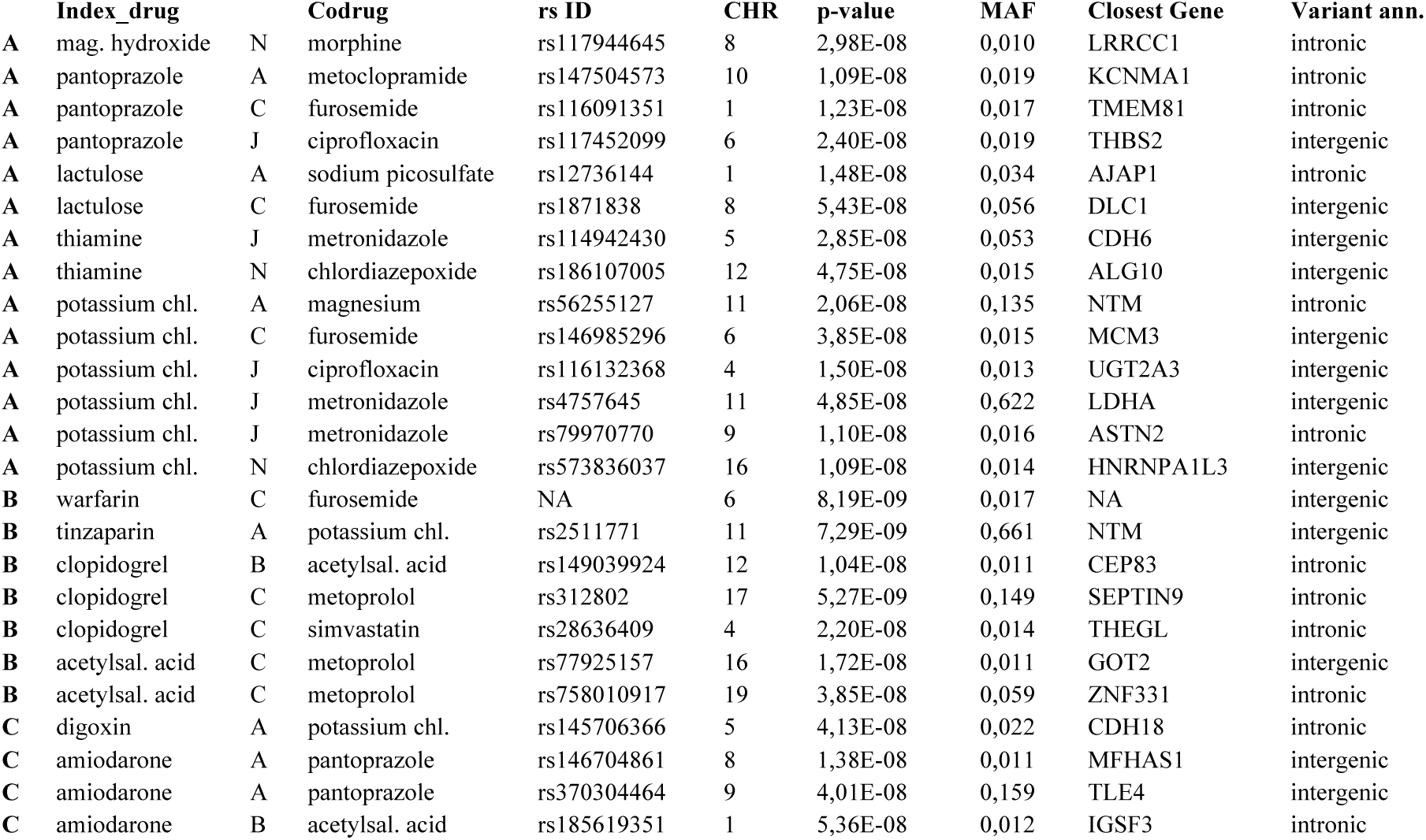
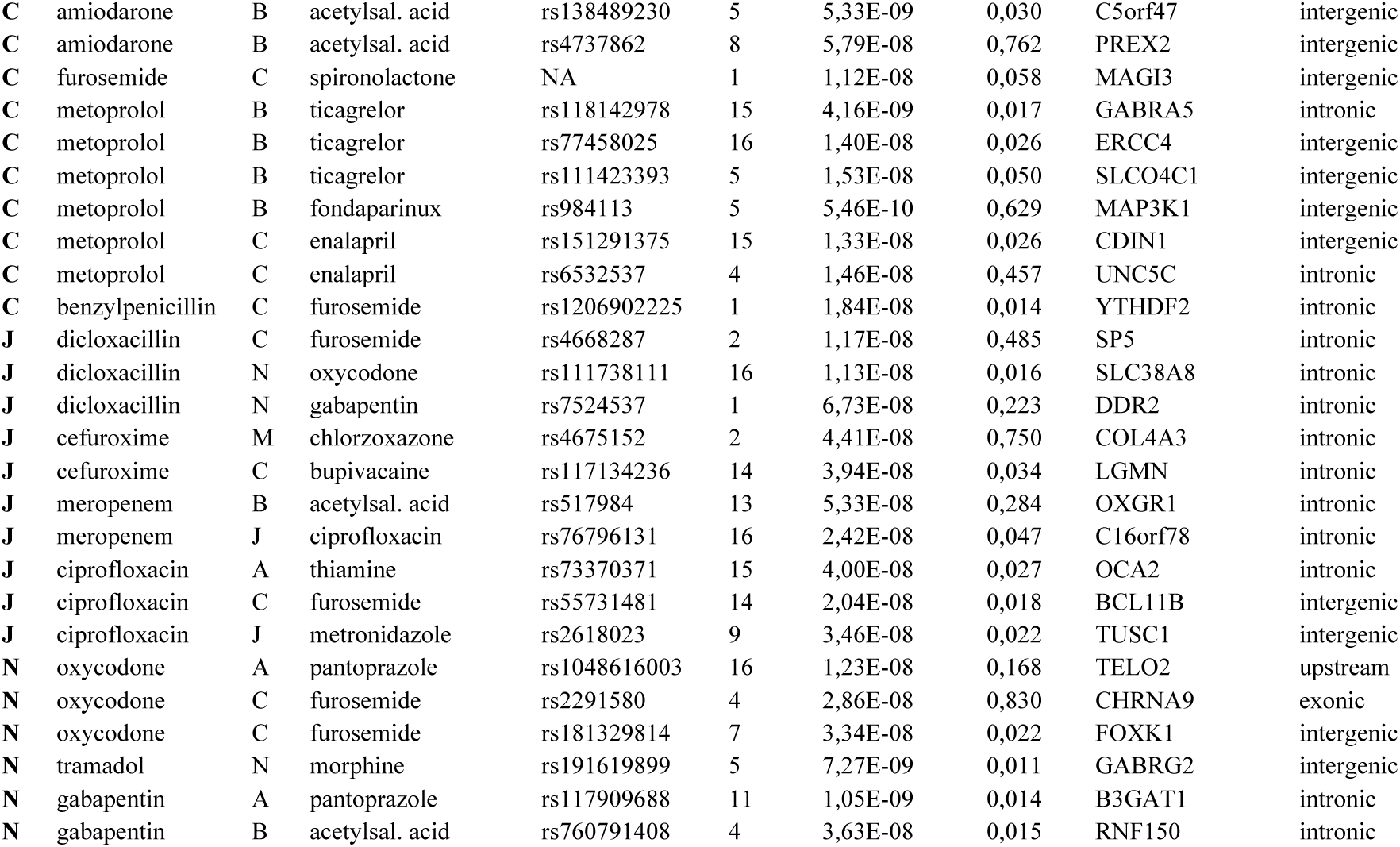
Genome-wide significant variants identified through the GWAS of 194 drug pair phenotypes. Bold letters in columns 1, and 3 indicate the main ATC group of each drug. **A** Alimentary tract and metabolism **B** Blood and blood forming organs **C** Cardiovascular system **J** Anti-infectives for systemic use **M** Musculo-skeletal system **N** Nervous system.

Across the 42 GWASs most showed little evidence of genomic inflation with lambda GC values between 0.9927–1.0802 (mean: 1.0302, SE: 0.0032) and LD-score regression intercepts between 0.9768–1.09 (SE: 0.0062–0.0084) (Table S4). Among the identified lead variants only two had previously been reported in any GWAS (OpenTargets^29^, GWAS Catalog^33^), namely rs984113 (metoprolol + fondaparinux) which has previously been found associated with breast cancer (<u>GCST004988</u>) and rs76796131 (meropenem + ciprofloxacin) which was associated with smoking initiation (<u>GCST007468</u>) (Table S6). We tested to see if a dosage change in any particular direction (increase or decrease) was driving the association with dosage changes for any of the lead variants. We found that for 25 of the lead variants, the association was driven by either dosage increase (16 variants) or dosage decrease (9 variants) of the index drug. For an additional 21 lead variants there was an association with both dosage increase and decrease, while the remaining five variants only showed an association with dosage change and not dosage increase or decrease (Table S5).

For each drug pair phenotype, we included patients admitted with different admission diagnoses, which could influence treatment regimens and the likelihood of dosage changes. To assess whether the admission diagnosis was directly associated with the variants, we performed chi-squared testing across the 51 significant variants. For 49 of them, we found no association while two variants (rs117944645 and rs4737862) had nominally significant associations (Table S7). However, none of these were significant after correcting for multiple testing (False Discovery Rate).

Gene-based testing showed significant associations (p < 4.0e-6) among 2 of the 42 drug pair phenotypes (Table S8). Four genes were identified, three of them (*DCLRE1C*, *MEIG1*, and *OLAH*) being associated with the benzylpenicillin + furosemide phenotype while the last gene (*TMPRSS2*) was associated with the ciprofloxacin + furosemide phenotype. There was no overlap between the genes identified through significantly associated variants and those identified through gene-based testing.

### Correlations among drug pair genetic effects

Although many of the 194 drug pair phenotypes shared index- or codrugs there was little correlation between the genetic effect sizes with the median Pearson correlation being 0.173 and 0.007 for pairs sharing the index drug and codrug, respectively. Only 59 of the in total 18,721 comparisons showed a correlation above 0.5 (min p-value = 2.02e-292) (Table S9). The majority of these, 50, came from comparisons of drug pair phenotypes with the same index drug. Among the comparisons with the highest correlations were drug pairs with the index drug clopidogrel, an antiplatelet agent, with mean correlation 0.543 (min p-value = 2.02e-292) and amiodarone, an antiarrhythmic agent, with mean correlation 0.517 (min p-value = 1.83e-144) (Figure S3). Several other index drugs also showed mean correlation above 0.5 but these were aggregated from only two drug pairs each and we did not investigate them further.

### Phenotype associations among lead SNPs

Using PheWAS we identified other phenotypes related to the 51 lead variants. PheWAS through OpenTargets identified 51 significant associations across 12 drug pair phenotypes (Table S10). We found a number of associations between variant rs4737862 (amiodarone: an antiarrhythmic agent + acetylsalicylic acid: used in pain/fever treatment) and counts of various white blood cell types as well as between variant rs6532537 (metoprolol: a beta blocker + enalapril: an ACE inhibitor prodrug) and a number of fat-related anthropometric phenotypes such (body fat percentage, trunk fat mass, etc.) The strongest association was seen between rs116091351 (pantoprazole: a proton pump inhibitor + furosemide: a diuretic) and mean platelet volume (p = 1,2E-20).

### Genetic associations through LD, expression, and regulation

The 51 lead variants passing genome-wide significance (p < 5e-08) were located mostly in non-coding regions (25, 24, 1, and 1 variants in intronic, intergenic, upstream, and exonic regions, respectively). We explored indirect mechanisms by which the lead variants may affect drug therapy. First, we tested whether any of the lead variants were in LD with deleterious SNPs (CADD score > 12.37) in the nearest gene. None of the lead SNPs were in LD (R^2 > 0.8) with any deleterious exonic variants in the nearest gene, thus making it unlikely that any putative genetic effects are being mediated through deleterious effects from these genes. Second, we investigated whether dosage changes could be mediated through regulation of gene expression. Using eQTL catalogue annotations we found that 3 of the 51 lead variants were significantly associated with the expression of, in total, 6 genes in at least one tissue (Table S11). Among the results, rs4757645 (potassium chloride + metronidazole) was associated with increased expression of *GTF2H1,* a transcription factor subunit, in 43 separate tissues including brain, muscle, and skin tissue. Third, we queried the lead variants in RegulomeDB, a database containing experimental support for regulatory variants ^34^. Eight variants received a RegulomeDB ranking of at least 1F indicating a high probability that the variants are involved in regulation (Table S12). A few interesting observations were also made for those variants with lower RegulomeDB rankings. For example, 7 and 11 variants intersected with chromatin immunoprecipitation (CHiP) peaks in the *CTCF* and *CEBPA* genes, both of which are well-characterized transcriptional regulators. Variant rs312802 (clopidogrel + metoprolol) intersected with CHiP peaks for the *CTCF* gene from more than 400 tissue samples. Lastly, analysis of differentially expressed genes (DEGs) in FUMA across all 57 genes showed a significant (FDR < 0.05) down-regulation in salivary gland tissue (Figure S4). Separate DEG analysis of genes identified from each individual drug pair phenotype did not identify any DEGs, likely because few genes were associated with each individual drug pair phenotype.

### Links between gene sets and disease phenotypes

We identified associations to 57 genes based on location, gene-based results, and eQTL associations (Table S13). Several of the 57 genes showed previous associations to relevant disease-related phenotypes. *SEPTIN9*, which was associated with the clopidogrel + metoprolol (an anti-platelet drug and a beta-blocker) phenotype, showed associations to blood- and cardiovascular related phenotypes such as “systolic blood pressure”, “cardiovascular disease”, and “hypertension” (OpenTargets references <u>GCST90025968</u>, <u>GCST007072</u>, and <u>GCST90038604</u>). Additionally, through OpenTargets’ ChEMBL annotations we found several small molecules used in pain management, including some analgesics, acting on *GABRG2* (associated with tramadol + morphine, both analgesics).

### Variant effects on primary care prescription-related phenotypes

Our GWASs used in-hospital data to define cases and controls. We speculated whether the identified lead variants would also affect primary care drug therapy in patients not admitted to a hospital. Using drug prescription data from DNPR covering patients not included in our GWAS analyses, we tested whether any of the 51 variants also associated with differences in drug therapy in primary care. We found no statistically significant differences in either number of prescriptions or prescribed dose for any of the variant-drug combinations (Table S14). Limiting the analysis only to periods where patients had been prescribed both drugs showed the same pattern.

## Discussion

We identified 51 different lead variants associated with dosage changes across 16 drug pair phenotypes. To our knowledge no other concurrent drug use GWAS has been reported to date. Given that only two of the 51 variants had previously been associated with a phenotype in any GWAS, this suggests that the concurrent drug use phenotype can help reveal genetic associations that may go unnoticed by studies of single-drug effects. Correlations among variant effect sizes between drug pairs were generally low, suggesting genetic backgrounds that may be more or less specific to individual drug pair combinations. This specificity may stem from the fact that many drugs act on several targets and can be metabolized by different proteins, meaning that each combination of drugs may affect a more unique set of molecular processes leading to differences in downstream effects^35^. Findings from studies of drug response in individual drugs also notice lack of overlapping loci between drugs in the same class, suggesting a high level of specificity in drug-effect pathways^36,37^.

For several of the drug pair phenotypes the associated genes could be linked to relevant disease phenotypes supporting the therapeutic relevance of those genes. For example, we found that *SEPTIN9* was associated with the clopidogrel + metoprolol drug pair phenotype. Clopidogrel is a platelet inhibitor used to decrease the risk of heart disease, while metoprolol is a beta-blocker used to treat high blood pressure. Previously *SEPTIN9* has been associated with hypertension. Our findings suggest that the association of *SEPTIN9* with those phenotypes may be mediated through drugs commonly used in treatment of those conditions^38,39^. While our findings do not provide any mechanistic insight, they may provide clues for outcome-affecting genes or pathways that go beyond those identified in more disease-centered approaches.

Previous research linked our lead variants to processes of expression and regulation. Of the 57 drug pair-associated genes, six genes were linked to three of the lead variants through eQTLs. For example, we found evidence of association between rs984113 (metoprolol: a beta-blocker + fondaparinux: an anticoagulant) and *SETD9*, which is involved in DNA methylation. Previous evidence has linked *SETD9* with both coronary artery disease (<u>GCST005196</u>) and blood cell distribution (<u>GCST90025988</u>) supporting *SETD9*’s involvement in cardiovascular disease. This suggests that differentiated regulation may affect disease phenotypes through processes of altered medication patterns. The general pattern of genetic regulation affecting drug therapy has also been suggested previously, through studies of enhancers, microRNAs and other regulatory elements ^40,41^. While few have studied genetic determinants underlying multi-drug treatment there is evidence pointing to such regulatory effects from specific drug groups such as anticoagulants^42,43^.

In our analyses we found no evidence of associations between dosage changes and variants located in the “usual suspects” of pharmacogenetically important genes such as Cytochrome P450 (*CYP*) genes. Many of these genes such as *CYP2C9* and *CYP2D6* are well-described and the clinical effects of their variability likewise^44,45^. Carrying a certain set of variants in one of these genes can classify patients into phenotypic groups such as “poor metabolizer” and “normal metabolizer”, phenotypes which have known effects on efficacy of common drugs^18^. Among the 194 drug pairs analyzed here, 99 included known metabolites of clinically actionable *CYP* genes, several with both drugs being known metabolites^46^. One explanation for their absence, may be that the pathways and mechanisms important during multiple drug use may be entirely different from those which are important during monotherapy. Many *CYP* genes are involved in drug metabolism, and the absence of these genes from our results, may suggest that more complex interactions, such as changes in distribution or absorption, may be involved. Therefore, we hope that future research will expand to cover not only monotherapy, but also therapy with multiple medications, which may lead to deeper mechanistic insights.

This work shows some of the opportunities that detailed, individual-level electronic patient record data, and especially data related to drug dosing, may hold for pharmacogenomic research. Biobanks and national registries provide unique opportunities for research made possible by the ability to link various molecular level data such as genomics and proteomics to deep and longitudinal medical records^47–50^. Here, we go a step further than most registry studies by looking at highly individualized in-hospital phenotypes that highlight specific changes in drug therapy. The used dosage data may be among the most accurate drug dosage measures that are systematically gathered for a large cohort. Still, this study comes with several limitations that may influence the transferability of our approach. First, dosage changes are not necessarily motivated by genetic variation. For certain drug pairs there may be clear clinical guidelines for changing the dosage, including cases in which the two drugs belong to the same drug class and/or have similar indications (e.g. tramadol and morphine). Second, differences in dosing regimens across diagnoses may affect the generalizability of our approach. Although none of the 51 significant variants displayed evidence of being associated with the admission diagnosis, we cannot exclude that this could be the case for certain specific diagnoses when using this approach. Additionally, patients with more severe disease, such as those receiving ciprofloxacin and meropenem for example, may experience different patterns of dosage changes depending on disease progression. Given the high prevalence of multimorbidity and polypharmacy in our patient population, fine-grained analysis is challenging, partly as some treatment groups may be quite small. Third, despite the hospital drug records generally being of high quality, there may still be ambiguities in the definitions used to define patient response. For example, patients that change from a specific dose of an index drug to a “pro re nata” dose are here also classified as having experienced a “change”, though they may still be taking the same dose.

Future research in personalized drug therapy may focus on several opportunities. For example, investigating how the direction and magnitude of dosage changes differ among patients may complement findings from the simpler “dosage change” phenotype. Such studies may also take advantage of more advanced modelling approaches that go beyond the simple binary phenotype classifications used here and in many other genetic studies. In our study we tried to capture effects covering many drugs across many different patients. However, more targeted approaches may benefit from more specific cohorts, tailored drug dosing definitions and greater statistical power, although the number of patients will be lower. Such targeted approaches may also be used to explore more complex phenotypes, such as drug-triads and drug-disease combinations, which could yield deeper insights into drug functionality and efficacy. Finally, it is important to establish evidence from diverse populations. Here, we had access to a somewhat homogenous Caucasian population, and we encourage others to complement our findings by studying populations that differ from ours in their representation of ancestry, age, and morbidities.

In conclusion, we linked in-hospital prescription records and genetic data from Danish registries for 117,814 Danish patients and identified 51 independent loci associated with changes in drug dosage during episodes of multiple drug use. We showed that several of these loci may be coupled to processes of gene regulation and expression in genes which, to our knowledge, have not previously been linked to drug therapy outcomes. Weak correlations between the genetic effects of similar drug pair phenotypes suggest distinct genetic backgrounds among drug pairs. We hope that our approach may be useful in learning more about effects of multi-drug therapy and eventually lead to better outcomes for patients.

## Data Availability

Summary statistics from the 42 drug pair phenotypes with significant variants will be made available via the GWAS Catalog. The data that support the findings of this study are available from the CHB, but restrictions apply to the availability of these data, which were used under license for the current study, and so are not publicly available. Data are however available with permission of the CHB steering committee and the Danish national scientific ethical committee.

## Declarations

### Authors contributions

A.P.H., H.C., and S.B wrote the manuscript; A.P.H., C.L.R., and S.B. designed the research; A.P.H., I.L., J.H.B., and C.L.R. performed the research; A.P.H., I.L., C.L.R., H.C., and M.H.Z. analyzed the data; E.B., G.M., E.S., C.M., O.B.P., H.U., S.R.O., K.B., M.S., T.F.H., H.B., C.E., K.M.D., L.Q., M.T.B., H.H., and S.B. contributed new reagents/analytical tools.

## Acknowledgements

We acknowledge all the members of the Brunak group, especially Timo Röder, Troels Siggaard, and Sedrah Balaganeshan without whom the study would not have been possible. Finally, we acknowledge the patients in the Copenhagen Hospital Biobank.

## Carbon footprint

We estimated the carbon footprint of computations from this project using CPU power usage estimates and carbon intensity measures from the Danish Energy Agency. The computations’ total carbon emission was estimated at **180.9 kg CO2eq**, equivalent to driving 1,940 km in a gasoline car. This figure only covers activities performed by the University of Copenhagen and not the genotyping and imputation performed at deCODE genetics.

## Abbreviations

ADE: Adverse drug event
ATC: Anatomical therapeutic chemical
ALT: Alternative allele
CHB: Copenhagen Hospital Biobank
COJO: Conditional Joint Analysis
CYP: Cytochrome P450
DBDS: Danish Blood Donor Study
DDI: Drug-drug interaction
DEG: Differentially expressed gene
DNPR: Danish national patient registry
eQTL: Expression quantitative trait loci
GWAS: Genome-wide association study
REF: Reference allele

