## Supplementary Figures S1-S5 for "Genome-wide associations spanning 194 in-hospital drug dosage change phenotypes highlight diverse genetic backgrounds in concurrent drug therapy"

### Additional file 1: Supplementary figures for Genome-wide associations spanning 194 in-hospital drug dosage change phenotypes highlight diverse genetic backgrounds in concurrent drug therapy

Alexander Pil Henriksen[1], Cristina Leal Rodríguez[2], Hannah Currant[1,3], Ioannis louloudis[1], Jorge Hernansanz Biel[1], Maria Herrero-Zazo[4], Ewan Birney[4], Thomas Folkmann Hansen[1], Gianluca Mazzoni[1], Amalie Dahl Haue[1,5], Henning Bundgaard[6,7], Christian Erikstrup[8,9], Khoa Manh Dinh[8,10], Liam Quinn[11], Mie Topholm Bruun[12], Henrik Hjalgrim[13,14], Erik Sørensen[10], Christina Mikkelsen[10], Michael Schwinn[10], Ole Birger Vestager Pedersen[7,11], Henrik Ullum[15], Sisse Rye Ostrowski[7,10], DBDS Genomic Consortium, Karina Banasik[16], Søren Brunak[1]

[1] Novo Nordisk Foundation Center for Protein Research, Faculty of Health and Medical Sciences, University of Copenhagen, Copenhagen, Denmark
[2] Copenhagen Prospective Studies on Asthma in Childhood, Copenhagen University Hospital, Gentofte, Denmark
[3] Nuffield Department for Population Health, University of Oxford, Oxford, UK
[4] European Molecular Biology Laboratory, European Bioinformatics Institute, Hinxton, UK
[5] Danish Headache Center, Department of Neurology, Copenhagen University Hospital, Rigshospitalet-Glostrup, Copenhagen, Denmark
[6] The Heart Center, Rigshospitalet, Copenhagen University Hospital, Copenhagen, Denmark
[7] Department of Clinical Medicine, Faculty of Health and Medical Sciences, University of Copenhagen, Copenhagen, Denmark
[8] Department of Clinical Immunology, Aarhus University Hospital, Aarhus, Denmark
[9] Department of Clinical Medicine, Health, Aarhus University, Aarhus, Denmark
[10] Department of Clinical Immunology, Copenhagen University Hospital - Rigshospitalet, Copenhagen, Denmark
[11] Department of Clinical Immunology, Zealand University Hospital, Køge, Denmark
[12] Department of Clinical Immunology, Odense University Hospital, Odense, Denmark
[13] Danish Cancer Society Research Center, Copenhagen, Denmark
[14] Department of Epidemiology Research, Statens Serum Institut, Copenhagen, Denmark
[15] Statens Serum Institut, Copenhagen, Denmark
[16] Department of Gynecology and Obstetrics, Copenhagen University Hospital Hvidovre, Copenhagen, Denmark

#### List of figures

Figure S1 – Flowchart of patient and drug pair filtering
Figure S2 – Graph showing the explained variance for each of the top 15 Principal Components
Figure S3 – Heatmap showing the pairwise beta correlations between drug pair phenotypes sharing index drug Clopidogrel or Amiodarone
Figure S4 – Bar chart from FUMA's website showing Differentially expressed genes in the 57 genes associated with drug dosage changes in this study
Figure S5 – Manhattan and QQ plots for all 42 drug pair phenotypes with at least one significant SNP

**Figure S1:** Flowchart of patient and drug pair filtering.

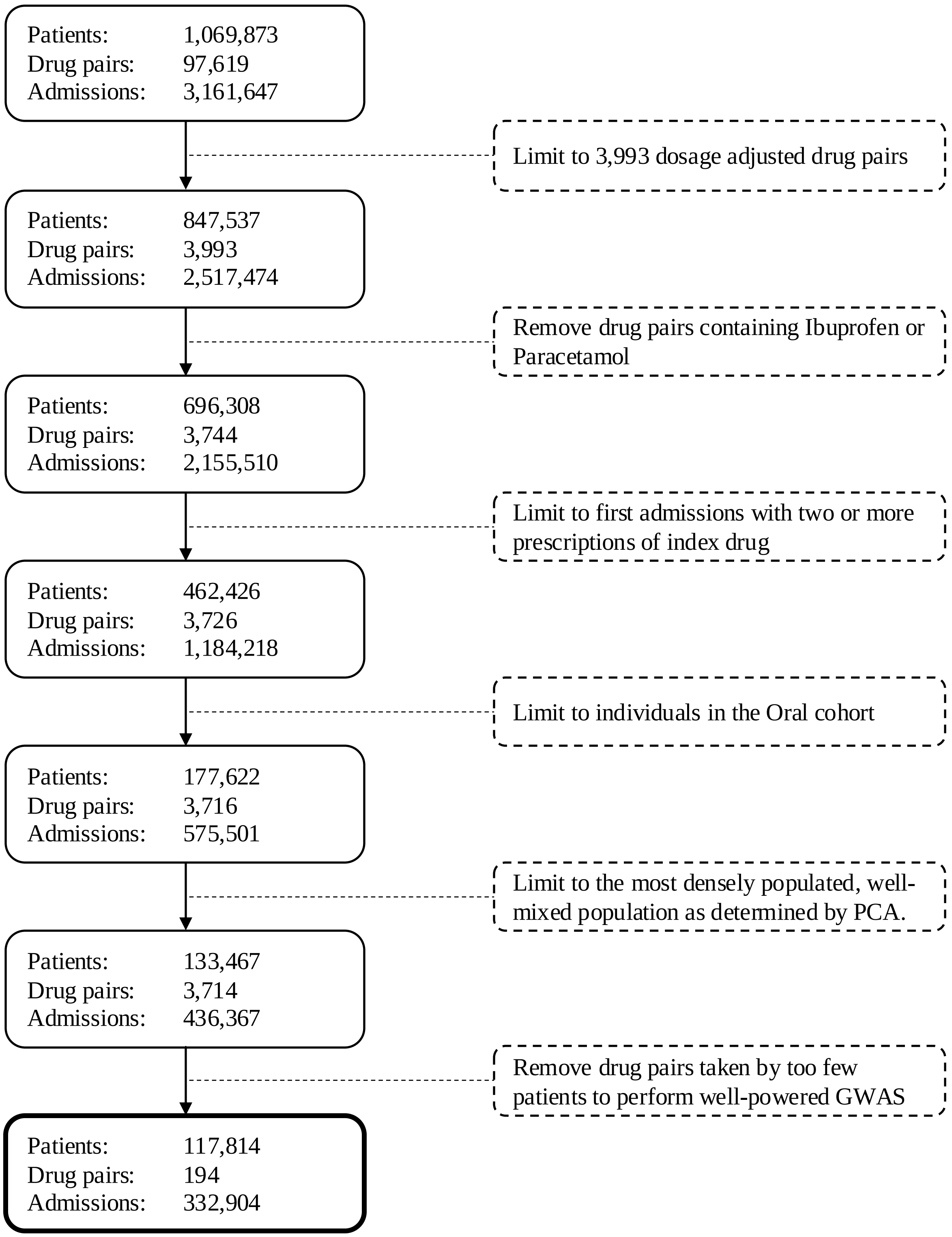

**Figure S2:** Proportion of variance explained by top 15 principal components across 133,467 genotyped patients.

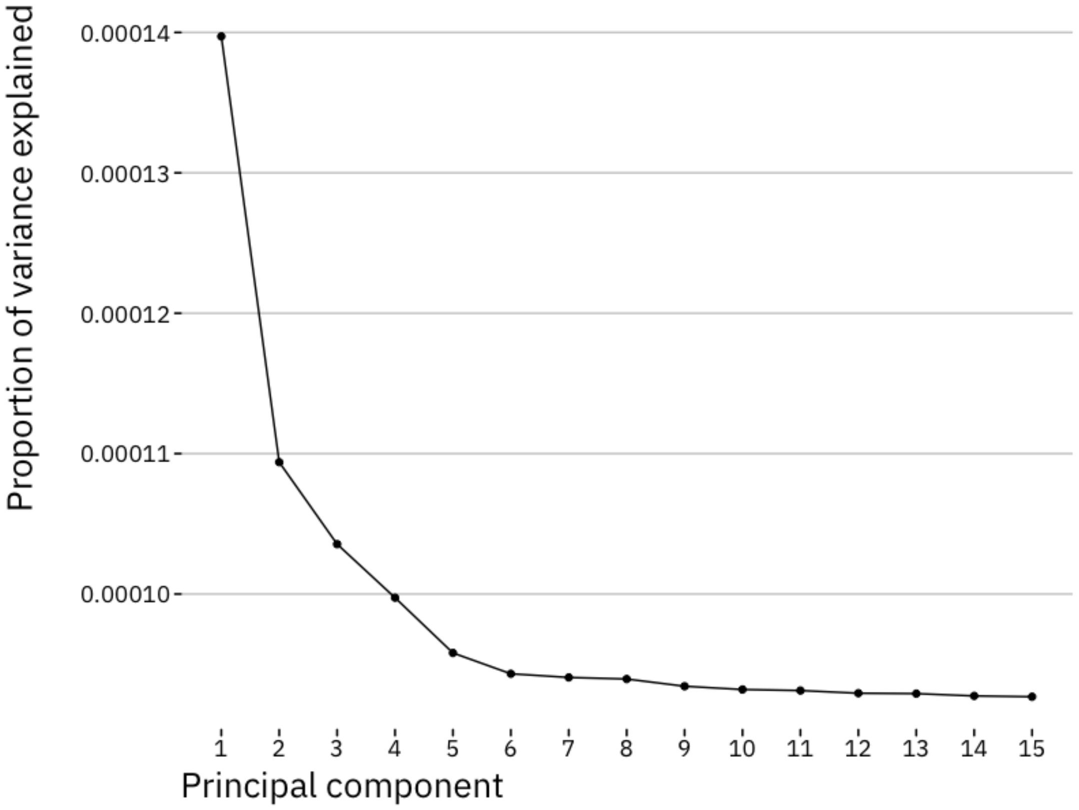

**Figure S3:** Correlation of variant effect sizes among drug pair phenotypes with shared index drugs. Any variant that with a p value lower than 5e-07 in any of the GWASs was included in the analsis. Effect sizes of the resulting 974 variants are compared across drug pair phenotypes. Here, only results for clopidogrel (ATC: B01AC04) and amiodarone (ATC: C01BD01) are shown, two drugs for which the drug pair phenotypes sharing that index drug showed a high level of correlation. The x- and y-axes shown indicate the codrug in the drug pair phenotypes that are compared.

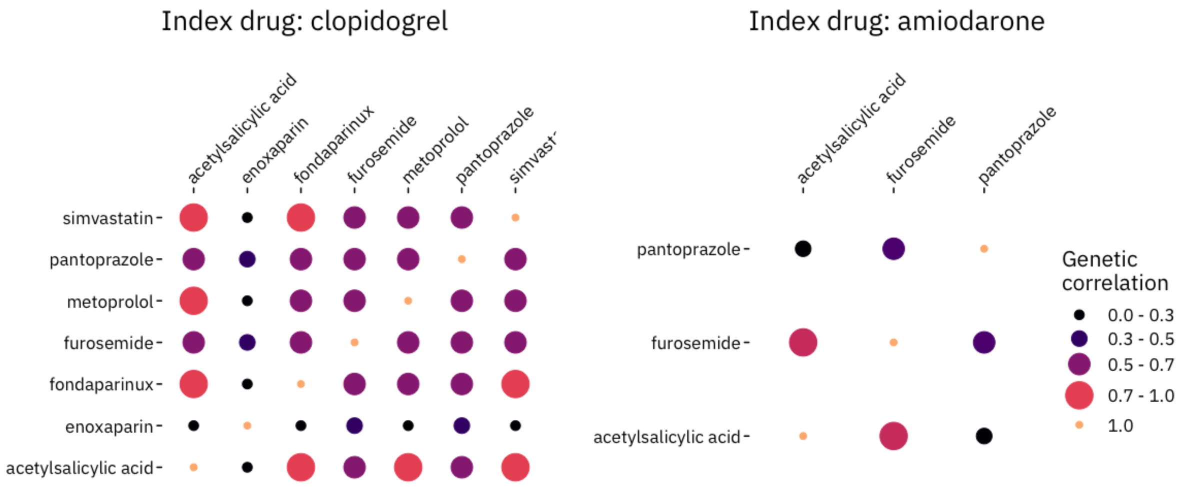

**Figure S4:** Tissue specificity of differentially expressed genes (DEGs) among 59 genes associated with the 42 drug pair phenotypes. Downloaded from FUMA website.

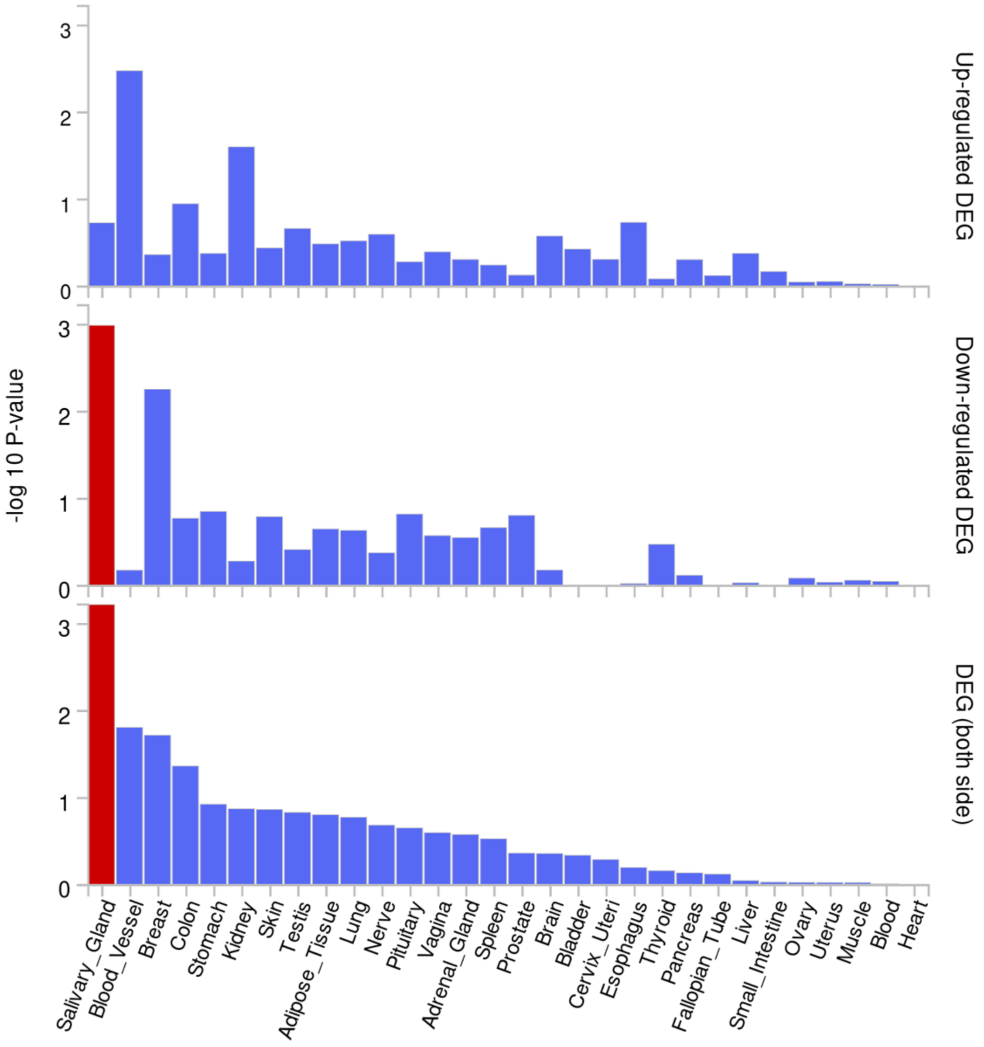

**Figure S5:** Manhattan- and QQ plots of all 42 drug pair phentoypes with at least one genome-wide significant

A02AA04_N02AA01: magnesium hydroxide + morphine

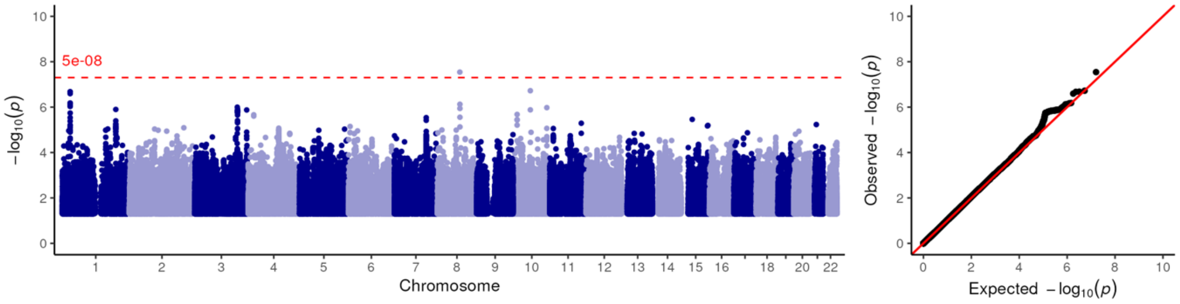


A02BC02_A03FA01: pantoprazole + metoclopramide

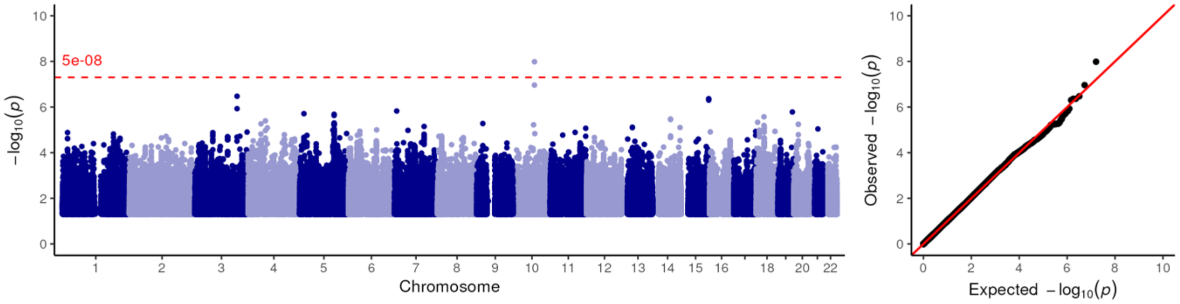


A02BC02_C03CA01: pantoprazole + furosemide

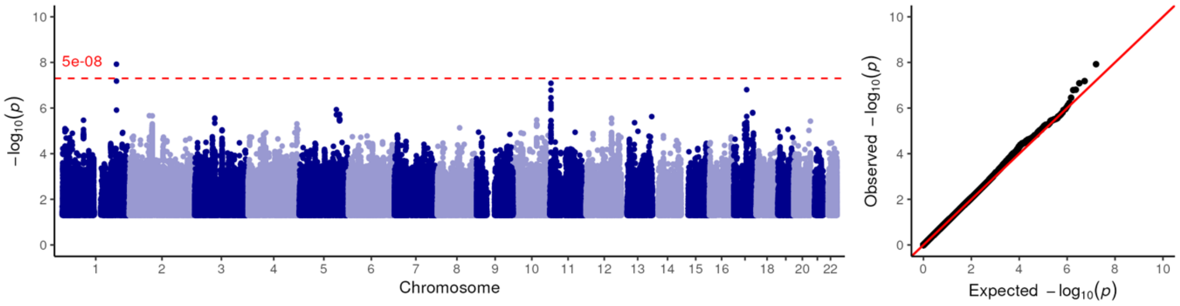


A02BC02_J01MA02: pantoprazole + ciprofloxacin

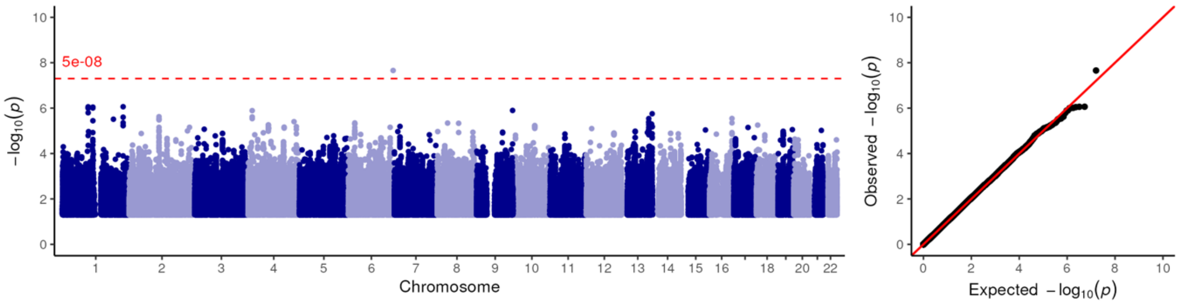

**Figure S5:** Manhattan- and QQ plots of all 42 drug pair phentoypes with at least one genome-wide significant

A06AD11_A06AB08: lactulose + sodium picosulfate

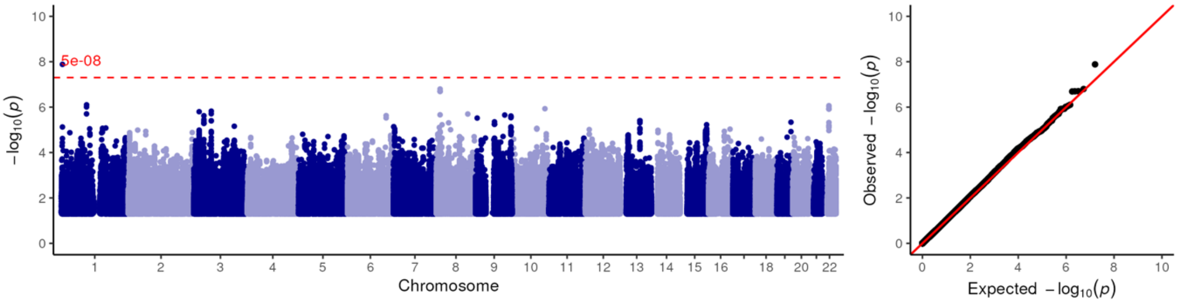


A06AD11_C03CA01: lactulose + furosemide

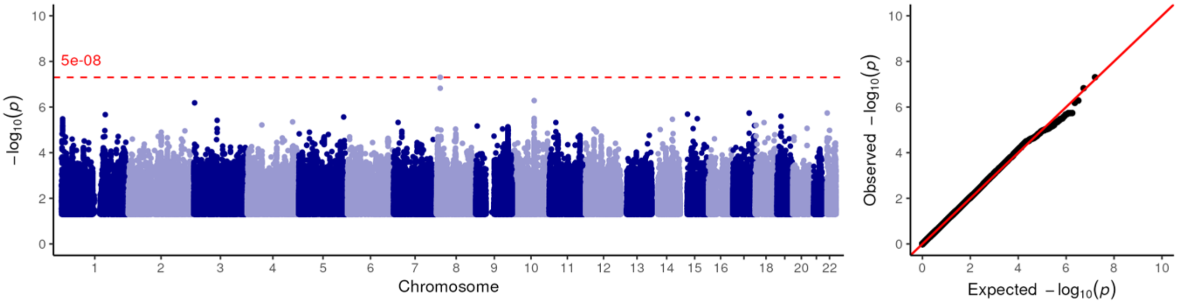


A11DA01_J01XD01: thiamine (vit B1) + metronidazole

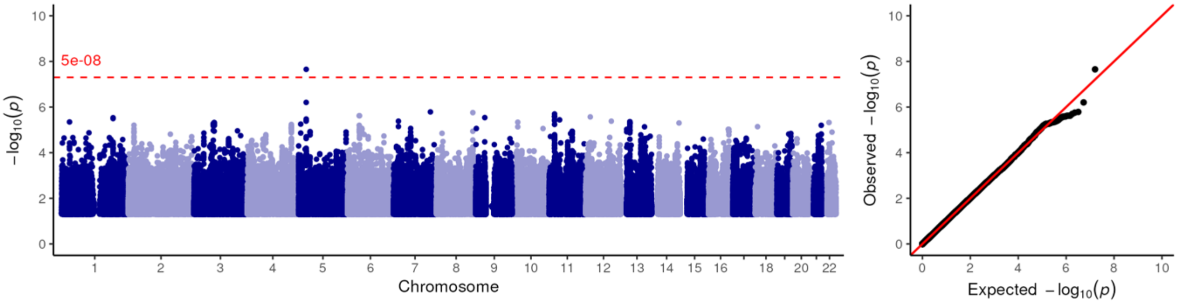


A11DA01_N05BA02: thiamine (vit B1) + chlordiazepoxide

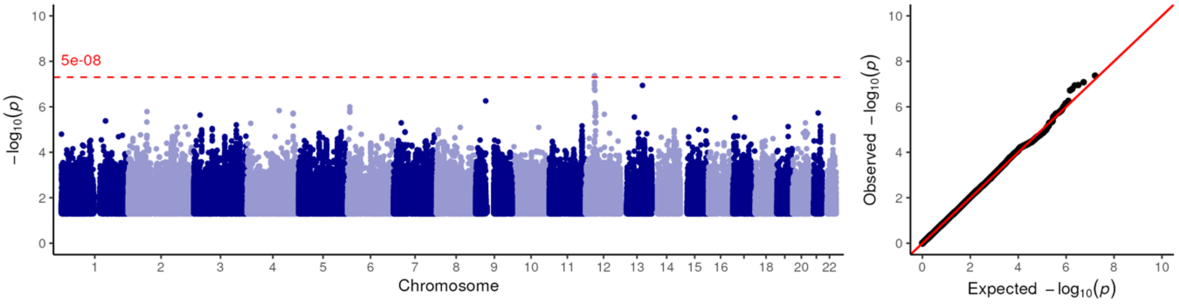

**Figure S5:** Manhattan- and QQ plots of all 42 drug pair phentoypes with at least one genome-wide significant

A12BA01_A12CC30: potassium chloride + magnesium (different salts in combination)

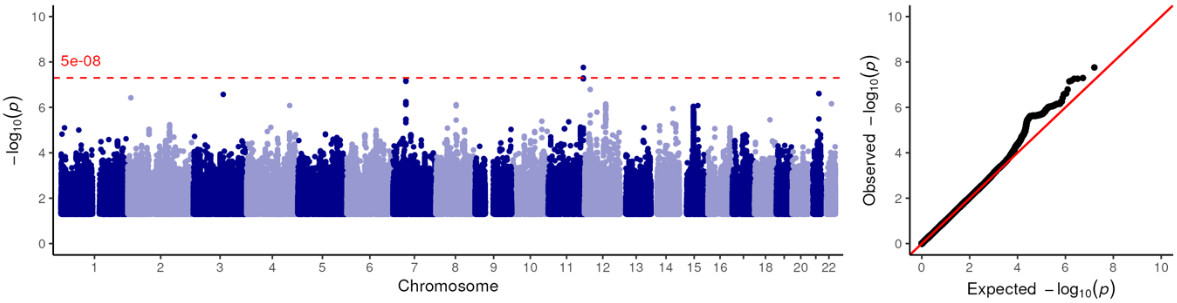


A12BA01_C03CA01: potassium chloride + furosemide

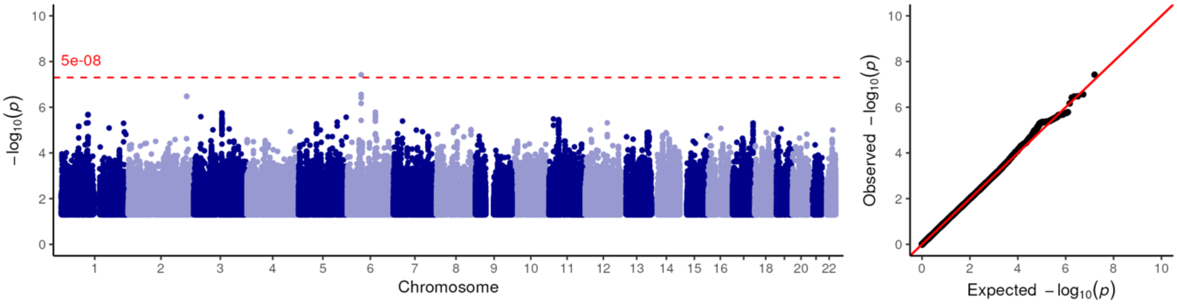


A12BA01_J01MA02: potassium chloride + ciprofloxacin

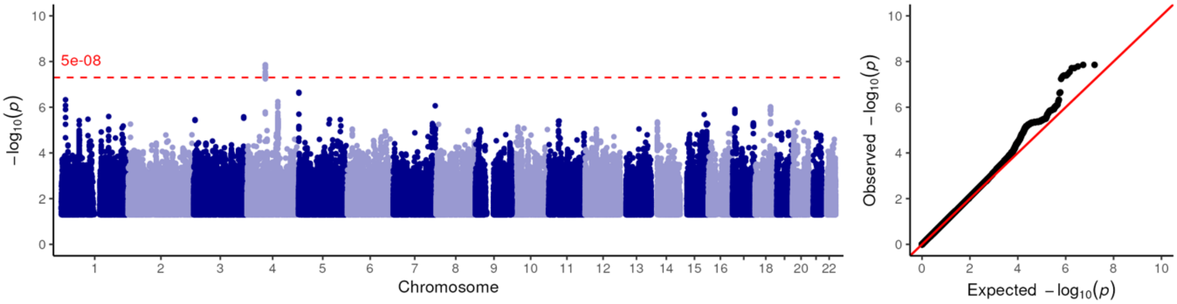


A12BA01_J01XD01: potassium chloride + metronidazole

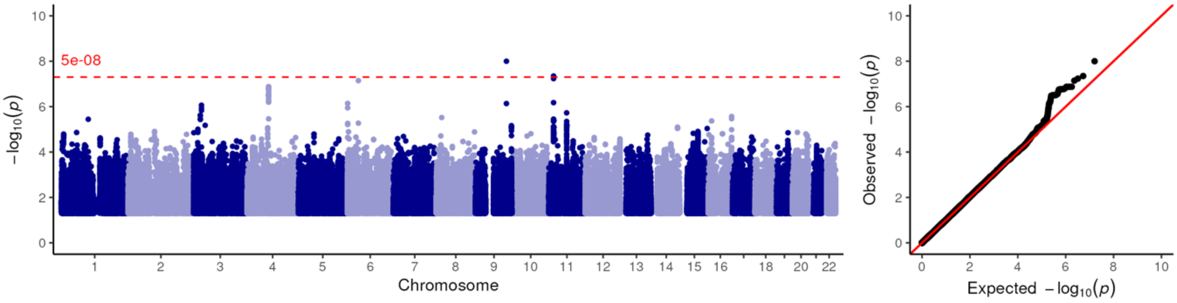

**Figure S5:** Manhattan- and QQ plots of all 42 drug pair phentoypes with at least one genome-wide significant

A12BA01_N05BA02: potassium chloride + chlordiazepoxide

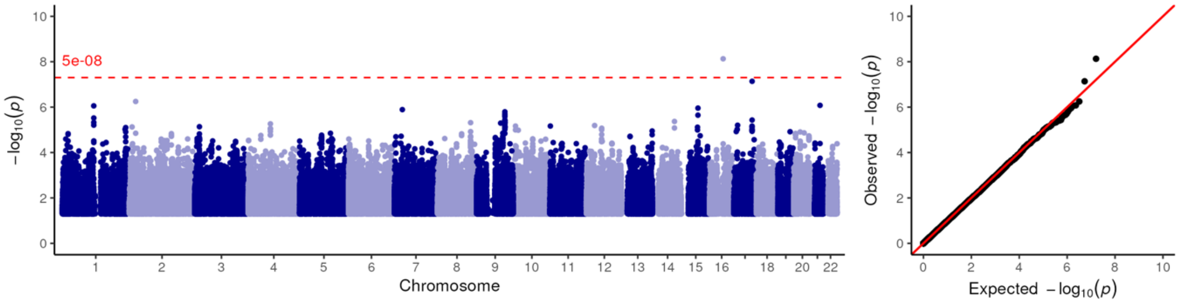


B01AA03_C03CA01: warfarin + furosemide

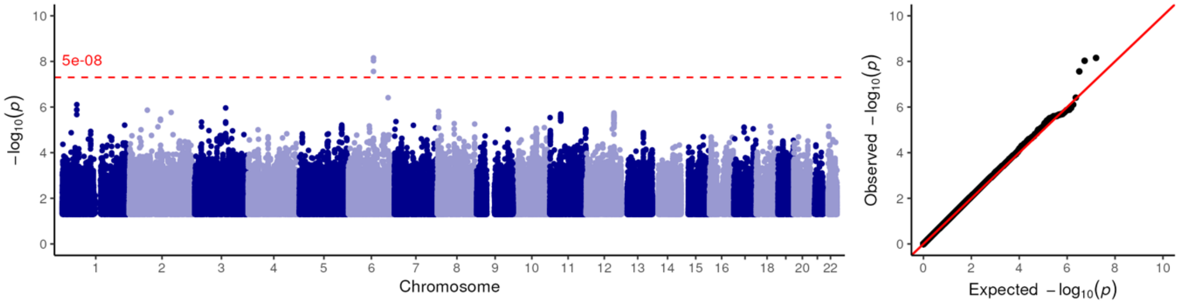


B01AB10_A12BA01: tinzaparin + potassium chloride

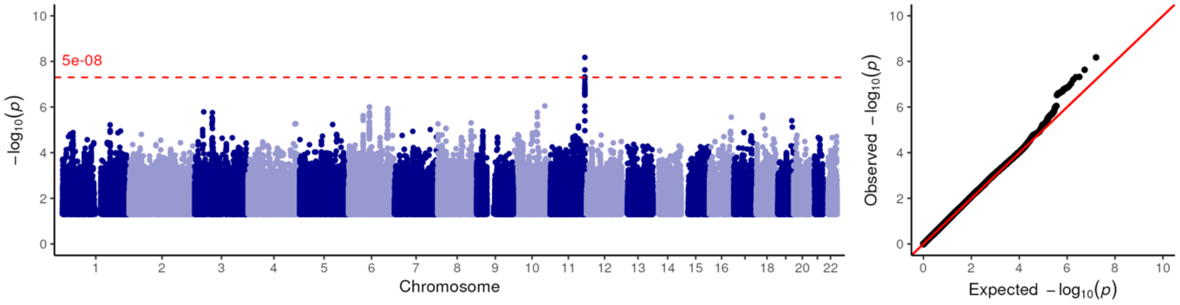


B01AC04_B01AC06: clopidogrel + acetylsalicylic acid

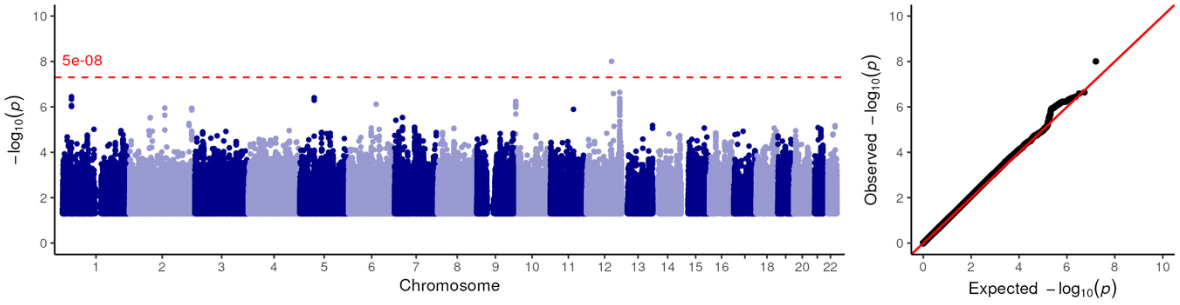

**Figure S5:** Manhattan- and QQ plots of all 42 drug pair phentoypes with at least one genome-wide significant

B01AC04_C07AB02: clopidogrel + metoprolol

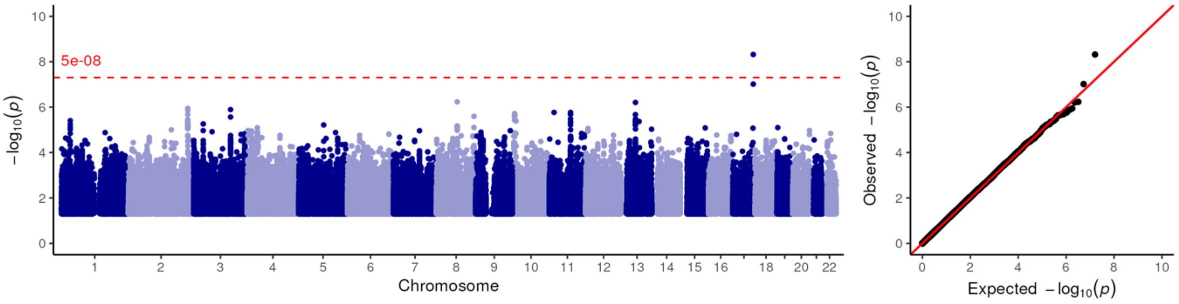


B01AC04_C10AA01: clopidogrel + simvastatin

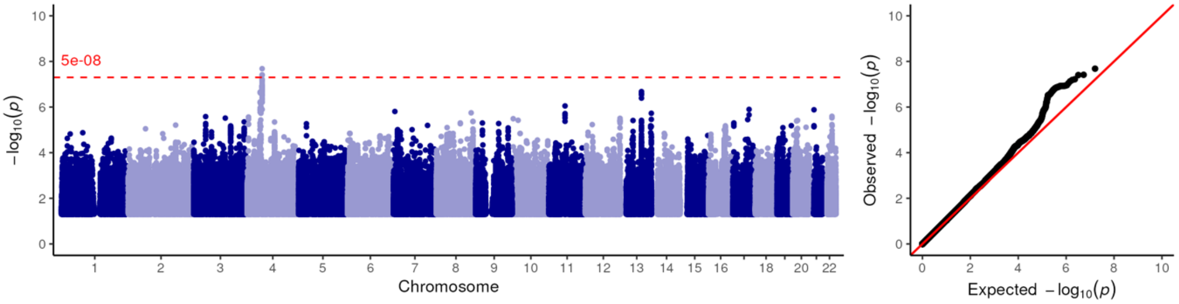


B01AC06_C07AB02: acetylsalicylic acid + metoprolol

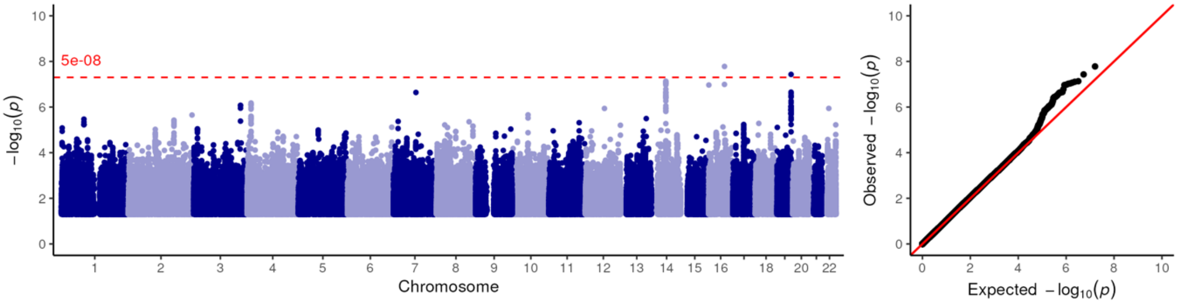


C01AA05_A12BA01: digoxin + potassium chloride

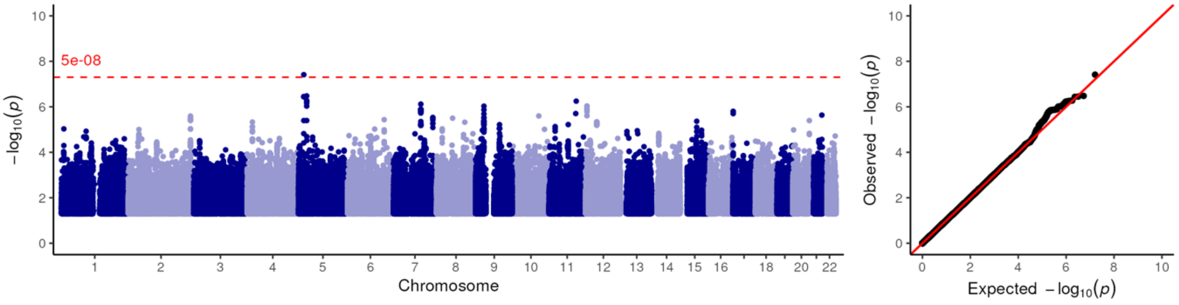

**Figure S5:** Manhattan- and QQ plots of all 42 drug pair phentoypes with at least one genome-wide significant

#### C01BD01_A02BC02: amiodarone + pantoprazole

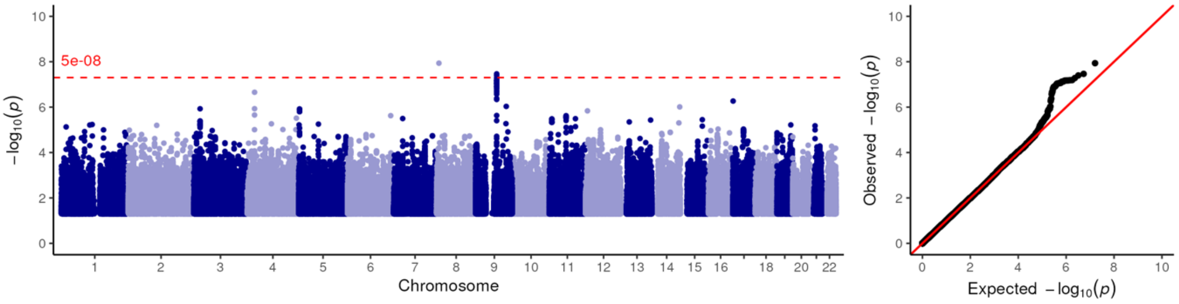


#### C01BD01_B01AC06: amiodarone + acetylsalicylic acid

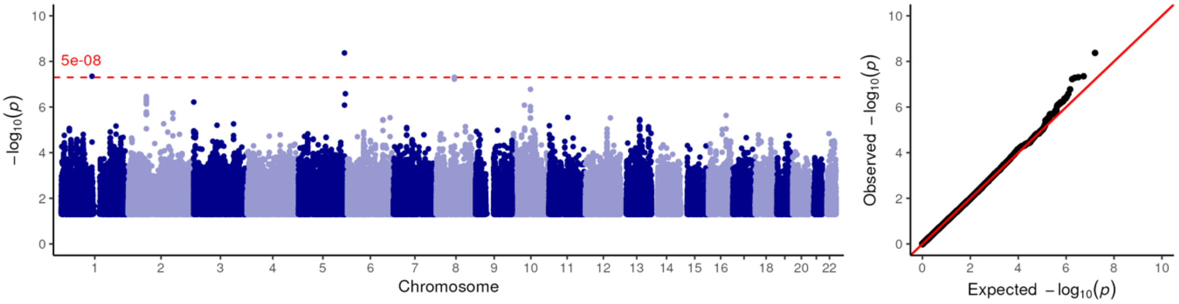


#### C03CA01_C03DA01: furosemide + spironolactone

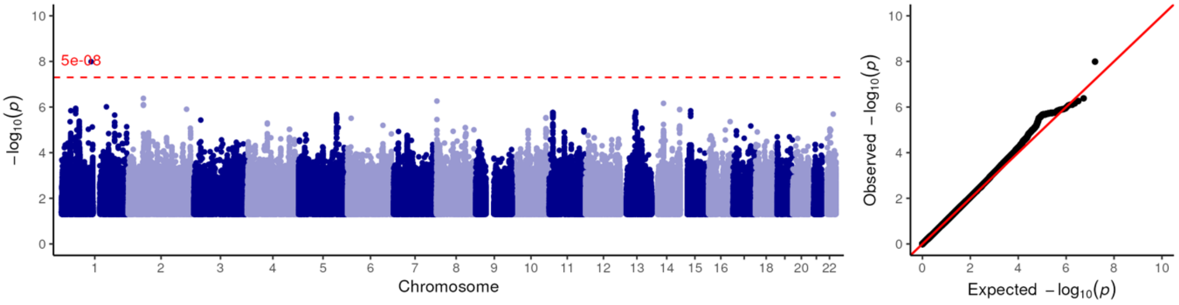


#### C07AB02_B01AC24: metoprolol + ticagrelor

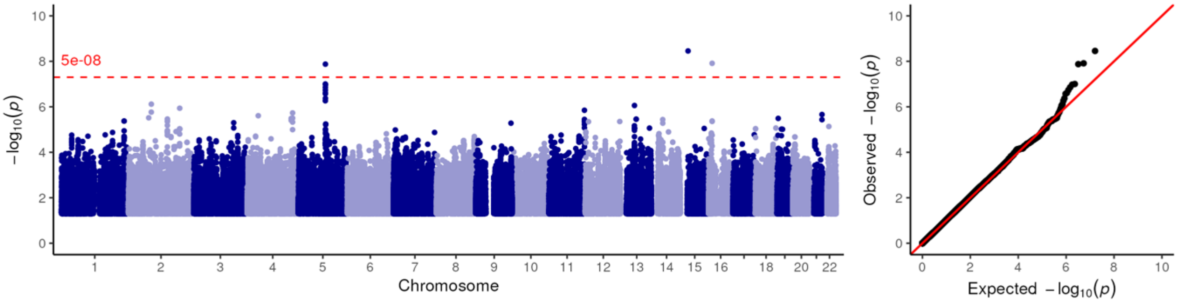

**Figure S5:** Manhattan- and QQ plots of all 42 drug pair phentoypes with at least one genome-wide significant

C07AB02_B01AX05: metoprolol + fondaparinux

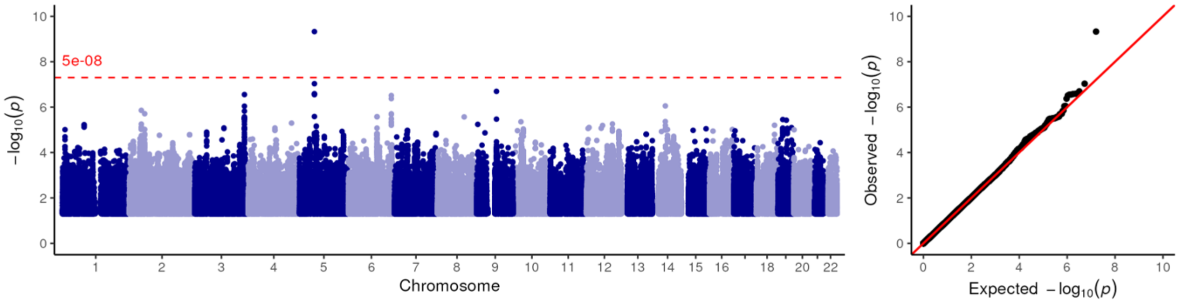


C07AB02_C09AA02: metoprolol + enalapril

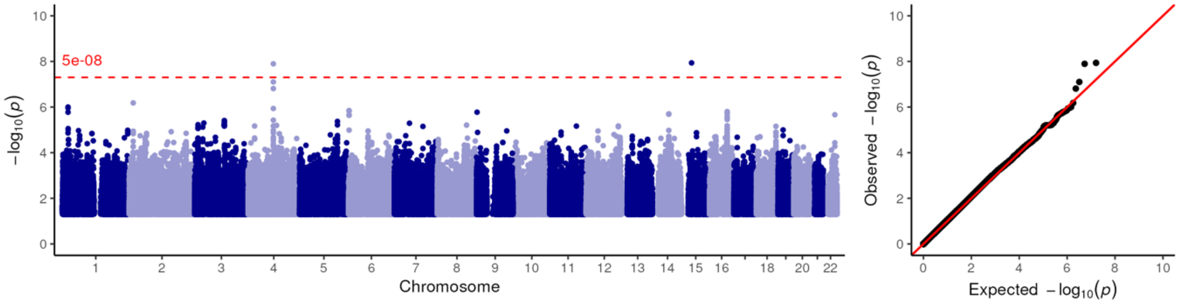


J01CE01_C03CA01: benzylpenicillin + furosemide

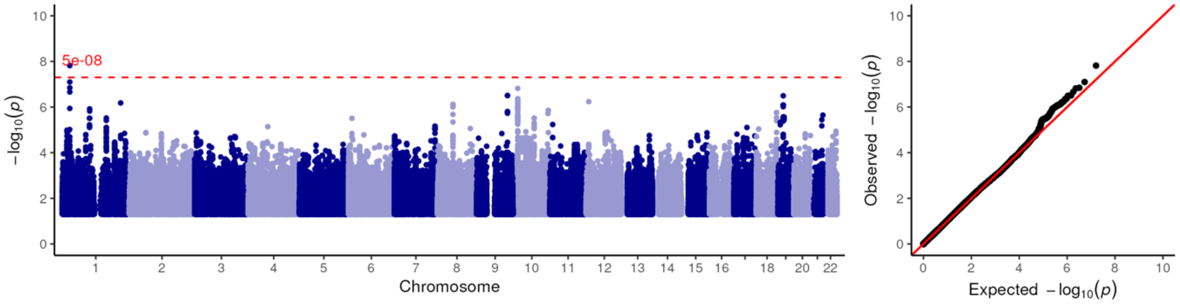


J01CF01_C03CA01: dicloxacillin + furosemide

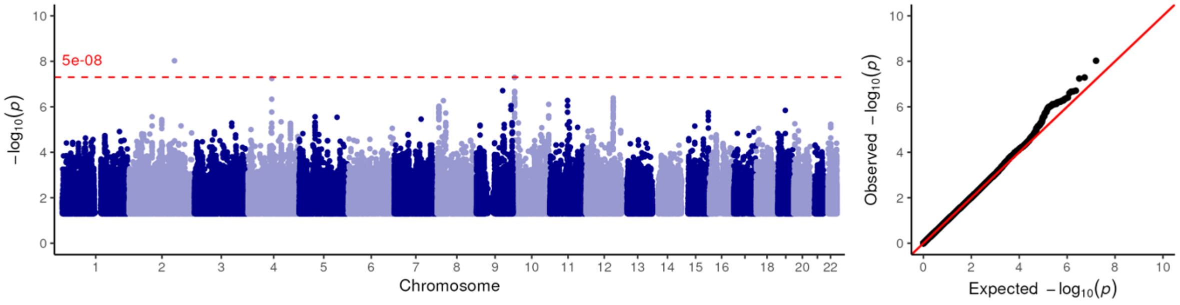

**Figure S5:** Manhattan- and QQ plots of all 42 drug pair phentoypes with at least one genome-wide significant

J01CF01_N02AA05: dicloxacillin + oxycodone

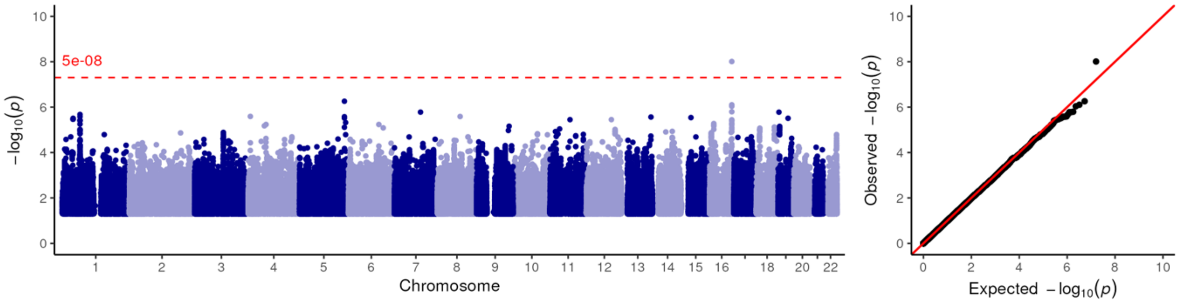


J01CF01_N03AX12: dicloxacillin + gabapentin

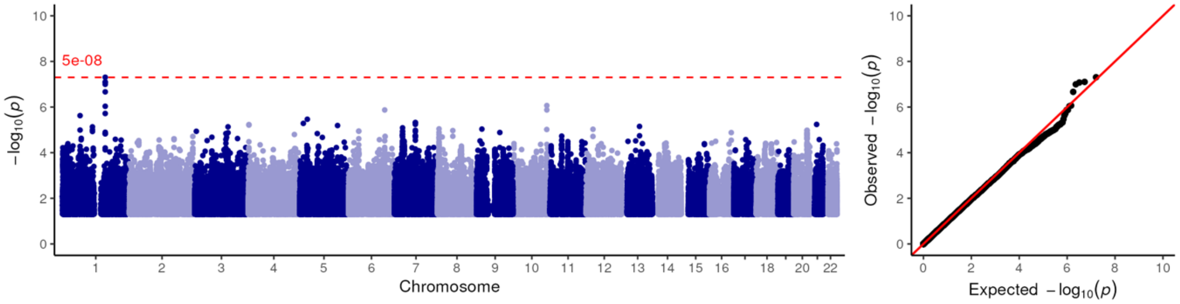


J01DC02_M03BB03: cefuroxime + chlorzoxazone

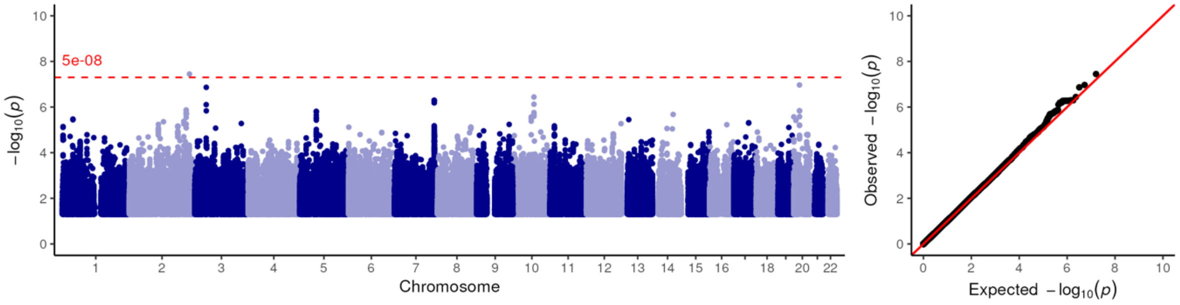


J01DC02_N01BB01: cefuroxime + bupivacaine

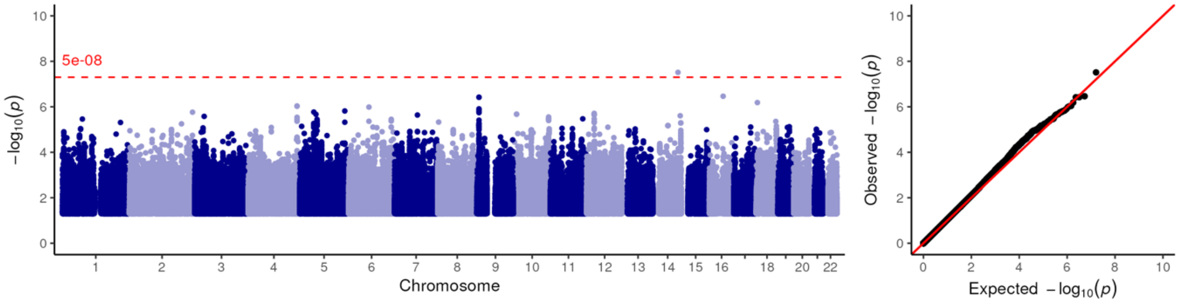

**Figure S5:** Manhattan- and QQ plots of all 42 drug pair phentoypes with at least one genome-wide significant

J01DH02_B01AC06: meropenem + acetylsalicylic acid

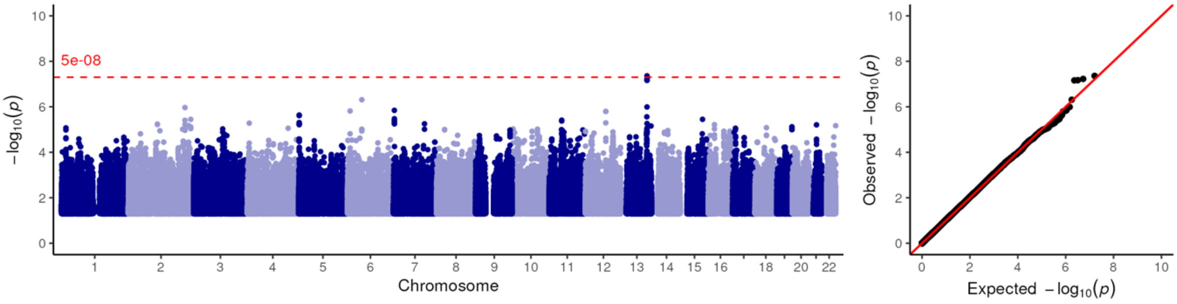


J01DH02_J01MA02: meropenem + ciprofloxacin

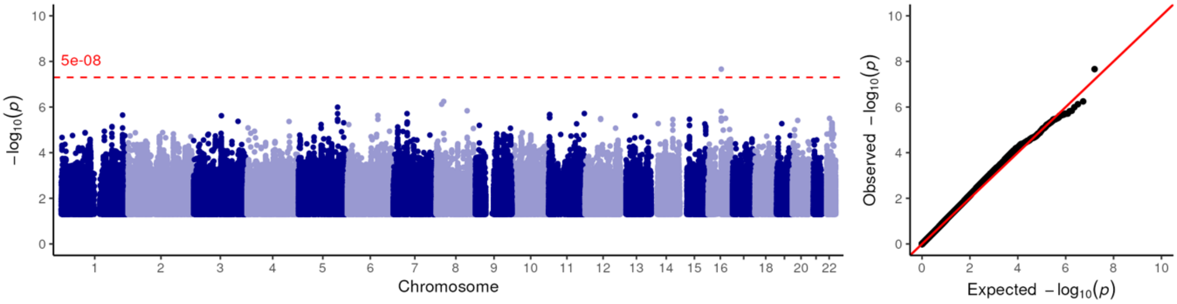


J01MA02_A11DA01: ciprofloxacin + thiamine (vit B1)

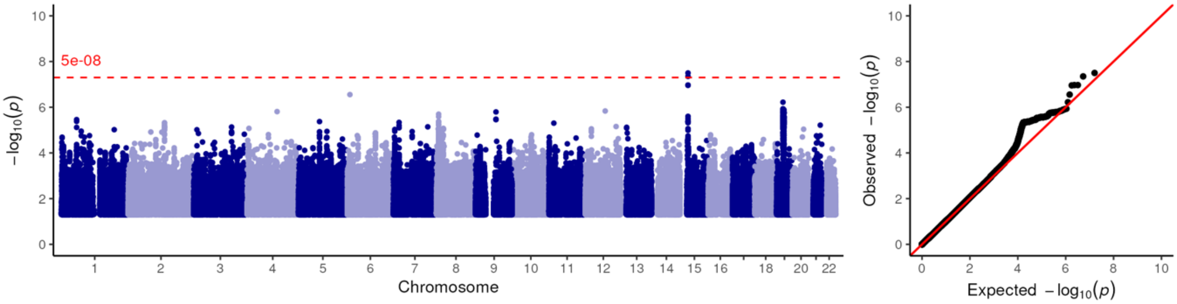


J01MA02_C03CA01: ciprofloxacin + furosemide

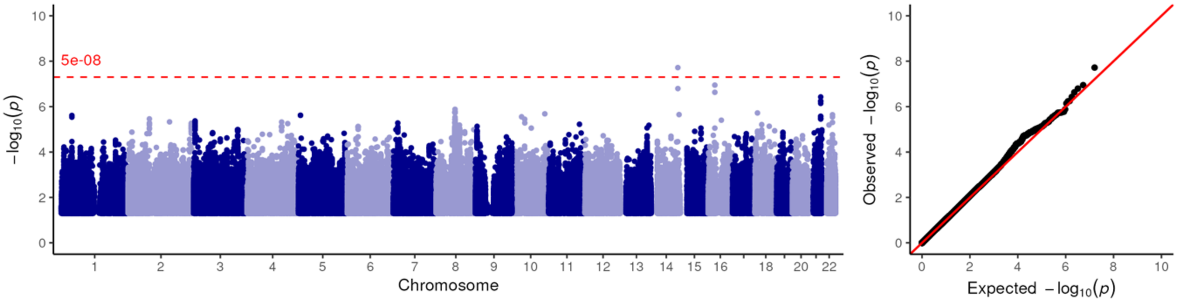

**Figure S5:** Manhattan- and QQ plots of all 42 drug pair phentoypes with at least one genome-wide significant

J01MA02_J01XD01: ciprofloxacin + metronidazole

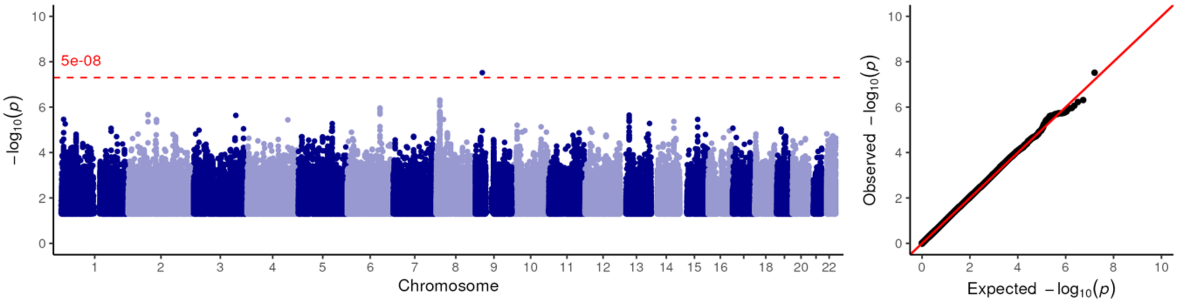


N02AA05_A02BC02: oxycodone + pantoprazole

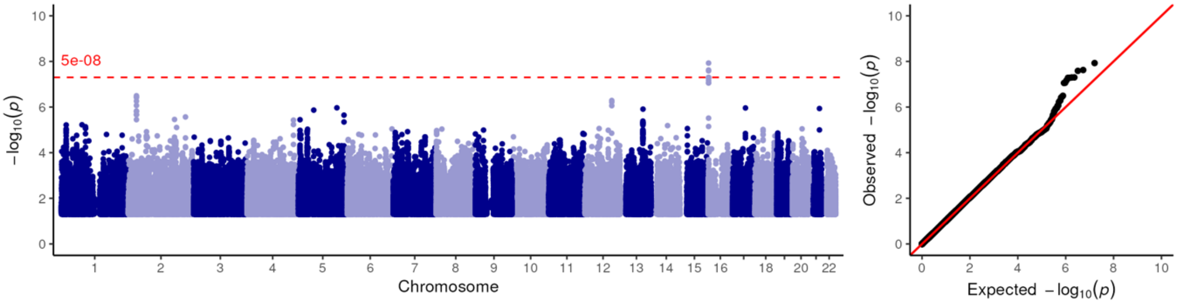


N02AA05_C03CA01: oxycodone + furosemide

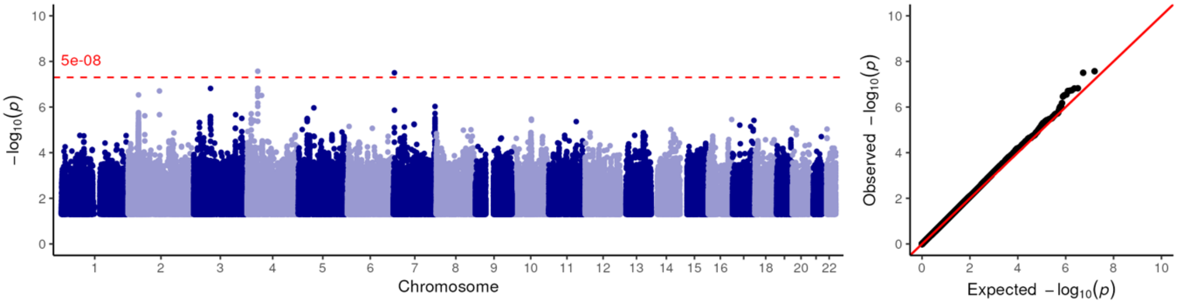


N02AX02_N02AA01: tramadol + morphine

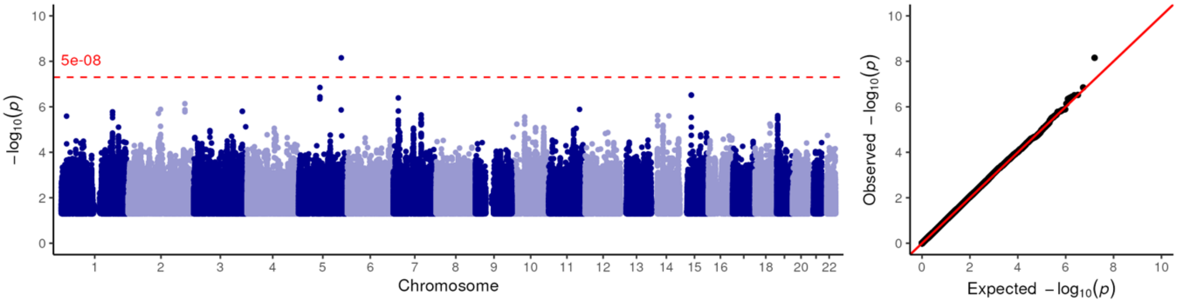

**Figure S5:** Manhattan- and QQ plots of all 42 drug pair phentoypes with at least one genome-wide significant

N03AX12_A02BC02: gabapentin + pantoprazole

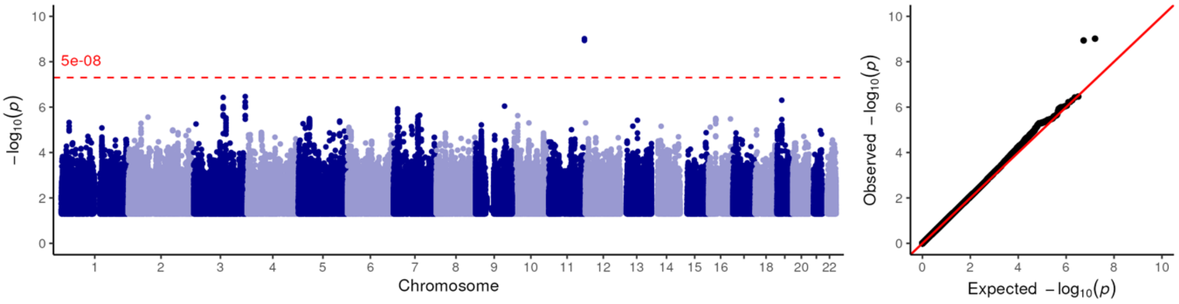


N03AX12_B01AC06: gabapentin + acetylsalicylic acid

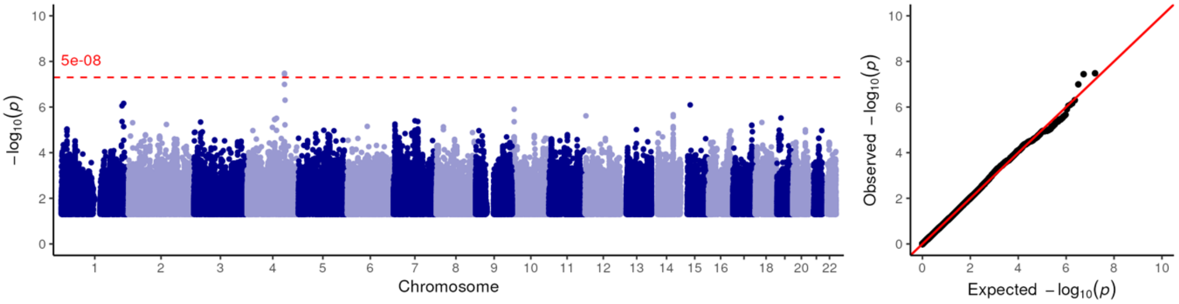
